# Modeling the synergistic interplay between malaria dynamics and economic growth

**DOI:** 10.1101/2023.12.09.23299755

**Authors:** Calistus N. Ngonghala, Hope Enright, Olivia Prosper, Ruijun Zhao

## Abstract

The mosquito-borne disease (malaria) imposes significant challenges on human health, healthcare systems, and economic growth/productivity in many countries. This study develops and analyzes a model to understand the interplay between malaria dynamics, economic growth, and transient events. It uncovers varied effects of malaria and economic parameters on model outcomes, highlighting the interdependence of the reproduction number (*R*_0_) on both malaria and economic factors, and a reciprocal relationship where malaria diminishes economic productivity, while higher economic output is associated with reduced malaria prevalence. This emphasizes the intricate interplay between malaria dynamics and socio-economic factors. The study offers insights into malaria control and underscores the significance of optimizing external aid allocation, especially favoring an even distribution strategy, with the most significant reduction observed in an equal monthly distribution strategy compared to longer distribution intervals. Furthermore, the study shows that controlling malaria in high mosquito biting areas with limited aid, low technology, inadequate treatment, or low economic investment is challenging. The model exhibits a backward bifurcation implying that sustainability of control and mitigation measures is essential even when *R*_0_ is slightly less than one. Additionally, there is a parameter regime for which long transients are feasible. Long transients are critical for predicting the behavior of dynamic systems and identifying factors influencing transitions; they reveal reservoirs of infection, vital for disease control. Policy recommendations for effective malaria control from the study include prioritizing sustained control measures, optimizing external aid allocation, and reducing mosquito biting.

## 1. Introduction

Malaria is a common source of human illness and death in many countries across the planet, especially in the World Health Organization (WHO) African Region, which harbored 95% of all the reported clinical cases of malaria and 96% of all the malaria-induced deaths in 2021 [1]. The health impact of malaria is particularly devastating among children under the age of five in the WHO African Region, with *≈* 80% of malaria-related deaths in 2021 occurring within this age group and region [1]. Despite progress in reducing the global burden of malaria (i.e., malaria-related morbidity and mortality) over the past decade, there has been an increase in cases and deaths, including 1 million additional cases from 2014 to 2015, 5 million cases from 2015 to 2016, and 13.4 million cases with 63,000 deaths between 2019 and 2021 [1, 2]. Interruptions in malaria control programs triggered by the COVID-19 pandemic were implicated for the increase in the global burden of malaria between 2019 and 2021. Since malaria is transmitted between humans by *Anopheles* mosquitoes, vector control measures including the use of traditional insecticide-treated nets (ITNs), piperonyl-butoxide (PBO) ITNs, indoor residual spraying, inhibiting the breeding of mosquitoes in proximity to human residences, etc., have been useful in combating malaria [3–6].

Apart from the substantial public health burden imposed by malaria to individuals, communities, and countries, the disease has significant economic consequences, including considerable direct healthcare costs, productivity losses, and barriers to economic development incurred by individuals, households, and governments in many countries in which it is endemic [7, 8]. Expenses related to prevention, diagnosis, treatment, hospitalization, and medications contribute to the substantial healthcare costs associated with malaria [9, 10]. Studies have shown that out-of-pocket expenditures for malaria-related services can push individuals and households further into poverty, exacerbating existing socioeconomic disparities [11–13]. Productivity losses resulting from malaria have a widespread impact, as infected individuals experience debilitating symptoms that hinder their ability to work or attend school. In particular, missed workdays, reduced productivity, and lower educational attainment are common outcomes [8, 14–19]. Additionally, malaria can cause agricultural workers to face challenges in tending to their crops or livestock, leading to decreased agricultural output and income [17–19]. The repercussions can be dire, leading to significant nutritional and economic hardships, particularly in resource-challenged rural communities that depend on subsistence agriculture and immediate natural resources for their livelihood. Malaria hampers economic development by deterring foreign investment, tourism, and trade [8, 20–22] since malaria-endemic regions are perceived as high-risk areas, discouraging potential investors and visitors. The presence of the disease inhibits industrial growth and limits opportunities for economic diversification, perpetuating a cycle of poverty and underdevelopment [20, 21, 23]. It should be mentioned that malaria has a profound impact on infants and children, and a significant proportion of malaria deaths occur in children under the age of 5. Expenditures on healthcare for malaria-afflicted children can significantly impact household income, while malaria-related absenteeism from school may hinder children’s future earning potential, contributing to a cycle of economic impact into adulthood [24]. Also malaria impacts adults, leading to economic losses and long-term health complications associated with chronic infections [7, 25, 26]. Studies have emphasized the economic strain imposed by malaria, including productivity loss among working-age adults, contributing to poverty in low-income areas [7, 27]. The disease correlates with significant reductions in annual economic growth [7, 28], and recent trends show an increase in global expenditures on malaria [13, 29]. Additionally, malaria ranks 19th in global disability-adjusted life years (DALYs) and 4th among infectious diseases in 2019 [30]. The economic impact of malaria extends beyond individuals, households, and countries, affecting regional and global economies. Healthcare costs and productivity losses drain national resources, diverting funds that could have been invested in critical sectors such as infrastructure and education. This collective burden hampers the ability of malaria-endemic countries to allocate resources effectively, impeding their overall economic progress. It is estimated that malaria costs the economy of African countries $12 million annually [31]. Addressing the economic impact of malaria requires comprehensive strategies that focus on reducing healthcare costs, improving productivity, and promoting economic diversification. By investing in malaria prevention, treatment, and control measures, countries can alleviate the economic burden, improve livelihoods, and foster sustainable economic growth.

The global health and economic trends of malaria underscore the need for more research into the efficacy and appropriate implementation of malaria control measures. One way to approach this is through mathematical modeling. Well-developed and calibrated mathematical models have played a critical role in understanding the dynamics of infectious diseases and in informing infectious disease control measures. In particular, much of the mathematical modeling literature on malaria and vector-borne diseases in general builds on the Ross-Macdonald framework for malaria of the 1900s [32, 33]. This basic but useful framework has been extended in various ways to account for more epidemiological and immunological aspects of malaria (e.g., [33–36]), demographic and feeding patterns of mosquitoes (e.g., [37–41]), environmental factors such as temperature (e.g., [42–46]), and various control and mitigation measures including the use of insecticide-treated nets and indoor residual spraying (e.g., [47–52])

Although these and other modeling efforts have made significant contributions to the study and control of malaria, an under-studied, yet crucial component to the success of malaria control programs is the dynamic feedback between the socio-economic landscape and malaria transmission. In particular, despite the overwhelming evidence that malaria and poverty are interconnected, and that malaria and other infectious diseases impact economic growth negatively [7, 8, 53], only a few mathematical frameworks attempt to explain this and the complex interplay between poverty and infectious diseases [54–56]. On the other hand, the study of transient dynamics in emerging, re-emerging, and endemic diseases has played an important role in improving disease management in real-time and understanding of patterns observed in epidemiological time-series [57]. These transients can emerge from endogenous and exogenous factors. In particular, the role of transients in a coupled malaria-economic system is of importance given the synergistic feedback between these two systems operating on different time scales and given the fact that although transient events occur within a relatively short timescale, they can have huge ripple global health and economic effects on such coupled system. Hence, a new mechanistic understanding of various processes leading to changes in malaria prevalence is key to identifying new interventions, understanding the intertwined relationship between malaria, socio-economic conditions, and transient events, and informing new empirical studies.

In this study, an epidemiological model that accounts for the dynamics of malaria, socio-economic features, and transient events is developed and analyzed. The framework is used to understand synergistic feedback between malaria dynamics and economic growth, as well as to assess the impact of transient events, particularly long transients on malaria dynamics and economic growth. To our knowledge, this is the first mathematical model for the transmission dynamics of malaria that accounts for all these factors in a single framework.

## 2. The model

### 2.1 Model formulation

In this section, an integrated model framework for the transmission dynamics of malaria that couples disease epidemiology with, human and mosquito population dynamics, as well as economic growth is developed. In the economic component of the framework, per capita economic yield or output (*y* = *f* (*k*) + *y*_*E*_, where *f* is a production function and *y*_*E*_ is external aid) is generated from labor (*L*) and per capita capital (*k*), which is defined as a stock resource that is used to produce goods and services. External aid for malaria involves financial and technical support from international organizations, governments, or non-governmental entities to combat and prevent malaria. This assistance includes funding, resources, and expertise to strengthen healthcare systems, implement preventive measures, and improve access to diagnostics and treatment. Examples include the Global Fund to Fight AIDS, Tuberculosis, and Malaria and the President’s Malaria Initiative, which provide aid for malaria control programs, bed net distribution, and drug procurement [58, 59]. As in [55], *f* is modeled through the Cobb-Douglas production function [60]. That is *f* (*k*) = *k*^*α*^(*AL/N*_*h*_)^1*−α*^, where *N*_*h*_ is the total human population, 0 *< α <* 1 is the capital share or elasticity coefficient, and *A* is technological progress or labor efficiency. It should be noted that per capita capital (yield) is given by *k* = *K/N*_*h*_ (*y* = *Y/N*_*h*_), where *K* (*Y*) is the aggregate capital (yield). Capital accumulates over time through savings of the unconsumed portion of the yield (1*− c*)*y* (where 0*≤ c≤* 1 is the factor for the consumed portion of the yield) at rate *r*, or depreciates at rate *σ* (see schematics in Fig. 1). Thus, in the tradition of [61, 62], the rate of change of capital is the difference between capital accumulation (*r*(1 *− c*)*y*) and capital depreciation (*σk*), i.e., 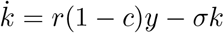 (last equation of Model (2.1)).

**Figure 1:**
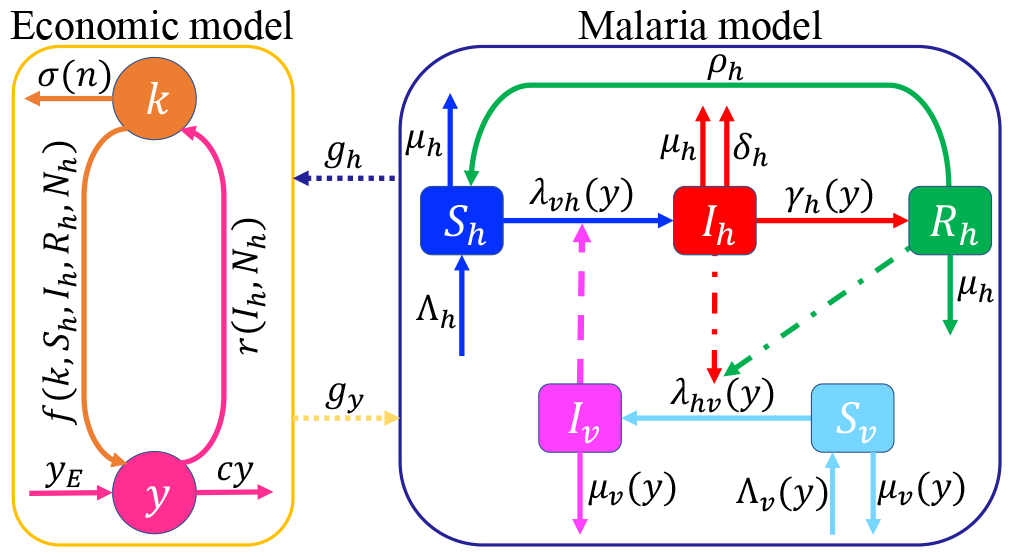
Schematics of the coupled economic-malaria model. The human population (*N*_*h*_) is split into susceptible (*S*_*h*_), infectious (*I*_*h*_), and partially immune (*R*_*h*_) individuals, while the mosquito population is split into susceptible (*S*_*v*_) and infectious (*I*_*v*_) individuals. Through production, capital (*k*) is transformed into yield (*y*), where a fraction of this yield (*cy*) is consumed, and the remaining portion ((1 *− c*)*y*) is reinvested into capital. External aid is denoted by *y*_*E*_. Dotted lines represent connections between the malaria and economic models. The malaria model is linked to the economic model through 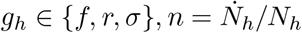 while the economic model is connected to the malaria model through *g*_*y*_ *∈* { *λ*_*vh*_, *λ*_*hv*_, *γ*_*h*_, Λ_*v*_, *µ*_*v*_}. Dash-dotted (dashed) lines indicate interactions between humans and mosquitoes leading to mosquito (human) infection. The rates and functional forms are described in the text.

In the malaria model, the total human population (*N*_*h*_) is divided into susceptible (*S*_*h*_), infectious (*I*_*h*_), and recovered or partially immune (*R*_*h*_) individuals. Similarly, the total mosquito population is divided into susceptible (*S*_*v*_) and infectious (*I*_*v*_) individuals. The model assumes no vertical transmission of malaria, so all human and mosquito births are into the susceptible classes at respective rates Λ_*h*_ and Λ_*v*_. The transmission dynamics of malaria involve the force of infection 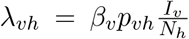 and 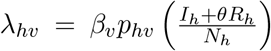, where *β*_*v*_ is the average number of bites a mosquito places on a human per unit time (commonly referred to as the human biting rate of mosquitoes), *p*_*vh*_ is the probability that an infectious mosquito infects a susceptible human, and *p*_*hv*_ is the probability that an infectious or partially immune human infects a susceptible mosquito. It is assumed that partially immune humans infect susceptible mosquitoes at a reduced rate depicted by the modification factor, 0*≤ θ ≤*1. Therefore, susceptible humans (mosquitoes) progress to the infectious human (mosquito) class at rate *λ*_*vh*_ (*λ*_*hv*_) after being infected. In other words new human and mosquito infections are given by *λ*_*vh*_*S*_*h*_ and *λ*_*hv*_*S*_*v*_, respectively. Humans (mosquitoes) from any of the human (mosquito) classes die naturally at per capita rate *µ*_*h*_ (*µ*_*v*_). Infectious humans either die from malaria at per capita rate *δ*_*h*_, or recover with partial immunity at per capita rate *γ*_*h*_. Given the relatively short lifespan of mosquitoes and the lack of empirical data supporting the effects of malaria on mosquito mortality, we assume that malaria does not directly cause mortality in mosquitoes. Also, since there is no empirical evidence of mosquito immunity to malaria, we assume that the infectious period for mosquitoes culminates with their death. Partially immune humans lose their immunity to become susceptible again at per capita rate *ρ*_*h*_.

The malaria model is coupled to the economic model through malaria-related medical costs (*ξI*_*h*_*y/N*_*h*_, 0*≤ ξ≤* 1) in the investment term, the human population growth rate 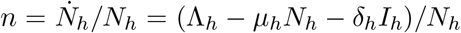 in the capital depreciation term, and the production function (*f* (*k, S*_*h*_, *I*_*h*_, *R*_*h*_, *N*_*h*_)). In particular, assuming that the total human population (*N*_*h*_) is proportional to the contribution of labor (*L*) to productivity and that clinically ill humans are less productive than their healthy counterparts, the production function (*f*) becomes 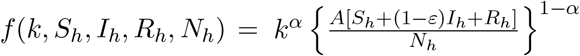, where 0 *≤ ε ≤* 1 is a modification parameter or factor to account for reduced productivity due to clinical malaria infection (Eq. (2.3)). It should be noted that *ε* = 0 (*ε* = 1) corresponds to a scenario in which clinically ill humans are fully (not) productive. Thus, the yield (*y*) is given by *y* = *f* (*k, S*_*h*_, *I*_*h*_, *R*_*h*_, *N*_*h*_)+*y*_*E*_. Conversely, the economic model is coupled to the malaria model through the mosquito biting rate (*β*_*v*_) in the forces of infection (*λ*_*vh*_ and *λ*_*hv*_ in Eq. (2.2)), the recovery rate from infection (*γ*_*h*_), the mosquito recruitment term (Λ_*v*_), and the mosquito mortality rate (*µ*_*v*_). Specifically, the biting rate of mosquitoes (*β*_*v*_) is modeled with the function 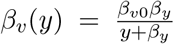, where *β*_*v*0_ is the maximum or background biting rate of mosquitoes (attained when *y→* 0) and *β*_*y*_ is a positive constant. Additionally, the human recovery rate is modeled through the functional form 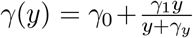, where *γ*_0_ is the background recovery rate (achieved when *y→* 0), *γ*_1_ is the additional economicdependent recovery rate, *γ*_*y*_ is a positive (half-saturation) constant, and *γ*_0_ + *γ*_1_ is the maximum recovery rate 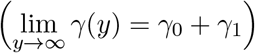. The mosquito recruitment rate (Λ_*v*_) is modeled with the function 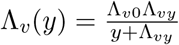, where Λ_*v*0_ is the maximum mosquito birth rate and Λ_*vy*_ is a constant. Furthermore, the mosquito natural mortality rate is modeled through the function 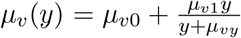, where *µ*_*v*0_ is the background mosquito mortality rate, *µ*_*v*1_ is an additional (economic-dependent) mortality rate, *µ*_*vy*_ is a half-saturation constant, and 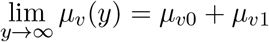.

A schematic representation of the coupled economic-malaria model is presented in Fig. 1, while the corresponding model system is given by Eqs. (2.1)-(2.3).

Using the schematics in Fig. 1 in conjunction with the detailed descriptions of variables and parameters in the text, we obtain the following coupled economic-malaria framework:

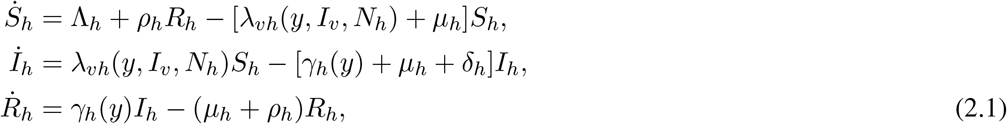

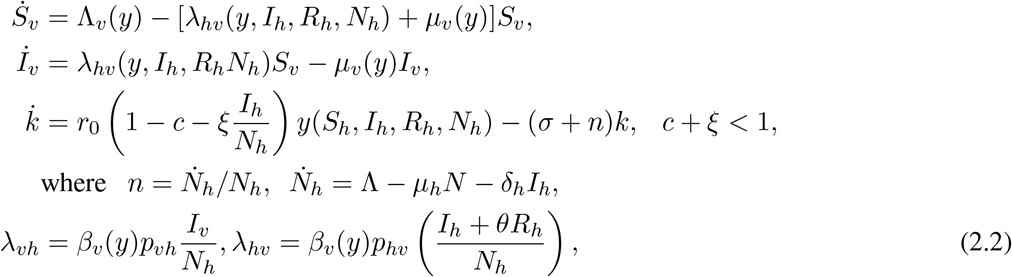

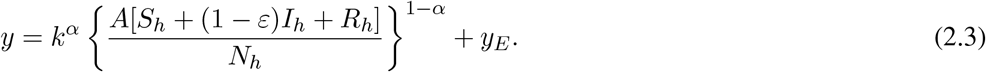

All model variables (*S*_*h*_, *I*_*h*_, *R*_*h*_, *S*_*v*_, *I*_*v*_, and *k*) are non-negative, as they correspond to human populations, mosquito populations, or physical quantities. Prescribing initial conditions of the form (*S*_*h*_(0), *I*_*h*_(0), *R*_*h*_(0), *S*_*v*_(0), *I*_*v*_(0), *k*(0)) = (*S*_*h*0_, *I*_*h*0_, *R*_*h*0_, *S*_*v*0_, *I*_*v*0_, *k*_0_), where *S*_*h*0_ *>* 0, *I*_*h*0_ *≥* 0, *R*_*h*0_ *≥* 0, *S*_*v*0_ *>* 0, *I*_*v*0_ *≥* 0, and *k*_0_ *>* 0 is necessary to fully define the model. Under these conditions, it can be easily verified that that the model (2.1) is well-posed from a mathematical, epidemiological and economic perspective. Furthermore, the region denoted by 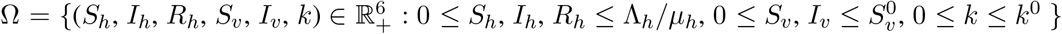, where 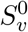 and *k*^0^ are the respective non-trivial disease-free equilibrium values of *S*_*v*_ and *k* is invariant and attracting for the system. Consequently, any solution trajectories originating within this region remain confined to it indefinitely.

### 2.2 The disease-free equilibrium and the basic reproduction number of the model

Using the specific functional forms for *γ, β*, Λ_*v*_ and *µ*_*v*_ described in the text, setting the right hand sides of Eqs. (2.1) as well as all disease states (*I*_*h*_, *R*_*h*_, and *I*_*v*_) to zero and solving for the non-disease states, we arrive at the disease-free equilibrium 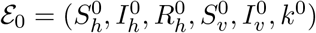, where

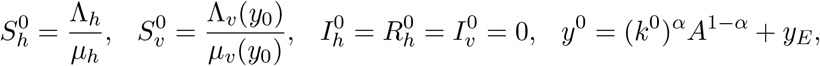

and *k*^0^ satisfies the equation *k*^0^*σ − r*_0_ (1 *− c*) [(*k*^0^)^*α*^*A*^1*−α*^ + *y*_*E*_] = 0, which can be rewritten as

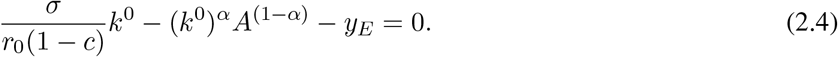

It is worth noting that the *k*^0^ equilibrium equation (Eq. (2.4)) can be solved in closed form if *y*_*E*_ = 0. Specifically, under the condition *yE*= 0, two closed form solutions arise: 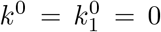 or 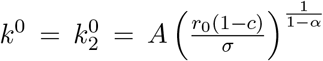

In general, the trivial disease-free equilibrium value of 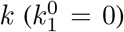 is unrealistic and unstable, while the positive non-trivial equilibrium value of 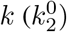 is locally asymptotically stable. For *y*_*E*_ *>* 0, the derivative of the left-hand side of (2.4) is *σ/*(*r*_0_(1 *− c*)) *− αA*^(1*−α*)^*/*(*k*^0^)^(1*−α*)^, which is negative for 0 *< k*^0^ *< A*(*σ/*(*αr*_0_(1*− c*)))^(1*−α*)^ and positive for *k*^0^ *> A*(*σ/*(*αr*_0_(1 *− c*)))^(1*−α*)^. Consequently, Eq. (2.4) has a unique positive solution (*k*^0^) when *y*_*E*_ *>* 0. This leads to the following result:

#### Lemma 2.1.

*Assume that α* ∈ (0, 1). *If y_E_ >* 0, *Eq*. (2.4) *has a unique positive solution. If y* = 0, *then* Eq. (2.4) *has two solutions:* 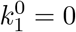 *and* 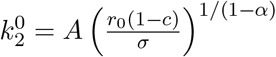, *and hence, the system has two disease-free equilibria*.

The local stability of the non-trivial disease-free equilibrium 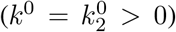 can be established using the Next Generation operator approach [63, 64]. This is a powerful tool for calculating the reproduction number (*R*_0_) of epidemiological models. Applying this approach yields the following reproduction number for the model (2.1):

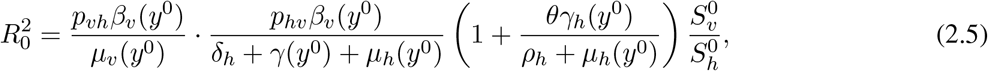

where 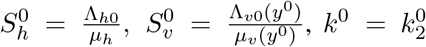 is the unique positive solution of the equilibrium equation (2.4), and *y*^0^ = *A*^1*−α*^*k*^*α*^ + *y*_*E*_ (*See Section S1 of the SI for details*). It can be verified that all conditions of the next generation operator approach are satisfied if the state variable, *k*, representing capital is treated as a disease-free state, irrespective of whether *y*_*E*_ = 0 or *y*_*E*_ *>* 0, provided that *k*^0^ is the positive solution to equation (2.4) (*See Section S1 of the SI for details*). This leads to the following result:

#### Theorem 2.2.

*Assume that α ∈* (0, 1). *If y*_*E*_ *>* 0, *all conditions of the next generation approach are satisfied if the state variable, k, representing capital, is treated as a disease-free state. If y*_*E*_ = 0, *all conditions of the next generation matrix approach are satisfied when the linearization is about the locally asymptotically stable disease-free equilibrium; that is, the DFE with* 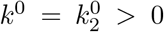. *In either case*, 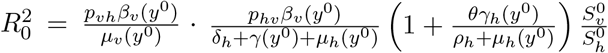, *where* 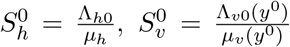 *is the unique positive solution of the equilibrium equation −k* (*σ*) *− r*_0_ (*c −* 1) (*A*^1*−α*^*k*^*α*^ + *y*_*E*_) = 0, *and y*^0^=*A*^1*−α*^*k*^*α*^+ *y*_*E*_.

*Proof. See Section S1 of the SI*.

**Remark 2.3**. *The value of α is often set to α* = 1*/*2 *in the standard Cobb-Douglas Production function [60]. This choice is based on empirical observations and mathematical convenience, and it implies that output is equally sensitive to changes in labor and capital inputs. Specifically, based on empirical fitting, α* = 1*/*2 *has been found to provide a good empirical fit for many industries and economies, where output growth appears to be roughly equally attributable to changes in labor and capital. For mathematical simplicity, the choice of α* = 1*/*2 *simplifies the mathematical properties of the production function, making it easier to analyze and derive results. Regarding other values of α, the choice of α may vary in different contexts, and alternative values are explored based on the specific characteristics of industries or economies under consideration [65, 66]*.

### 2.3 Endemic equilibria

The existence of endemic equilibria for the model system (2.1) is studied for the special case in which is 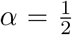.

Suppose an endemic equilibrium of the model (2.1) is denoted by 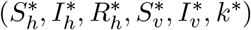, Let 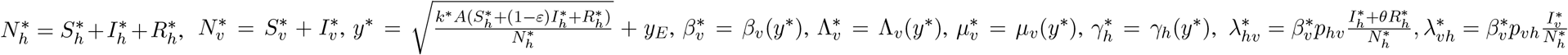. Then, at equilibrium,

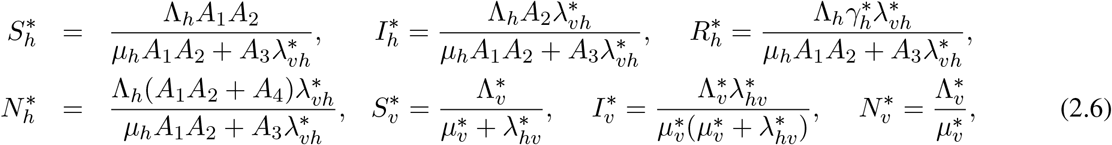

where

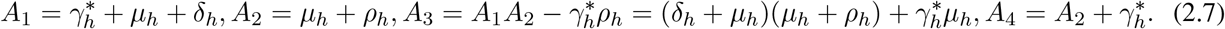

By substituting the equilibrium values of *I*_*h*_, *R*_*h*_, and *N*_*h*_ into the expression for *λ*_*hv*_, and similarly substituting the equilibrium values of *I*_*v*_ and *N*_*h*_ into the expression for *λ*_*vh*_, and then proceeding with the established methodology for variable elimination, we arrive at the ensuing equation for 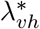

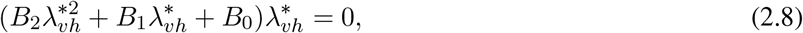

Where

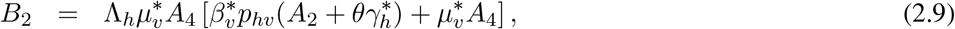

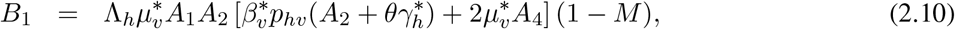

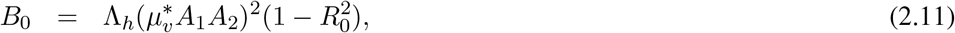

Where

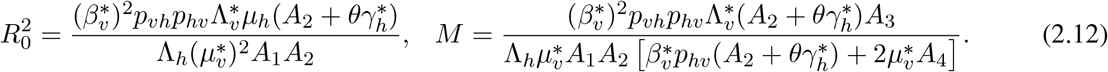

For any given *y*^*\**^, the endemic equilibrium can be found by solving Eq. (2.8), which gives 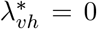 and 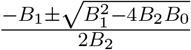. The case in which 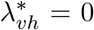 corresponds to the disease-free equilibrium. The possible number of positive roots and hence endemic equilibria (0, 1, or 2) determined by the signs of *M* and 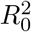 are summarized in Table S2 of the SI. Comparing the expressions of 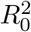 and *M*, the inequality 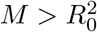 is true if and only if

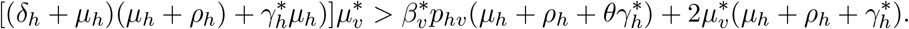

Specifically, this inequality holds when the disease-induced mortality rate (*δ*_*h*_) exceeds some threshold. This observation aligns with findings from other investigations, as exemplified by studies in [49, 50], where a backward bifurcation emerges with an increase in the disease-induced death rate beyond a certain threshold. It should be underscored that while the inequality may be satisfied through variations in other parameters, the choice of *δ*_*h*_ is particularly motivated by its role as a contributor to backward bifurcations [49, 50, 67]. Setting 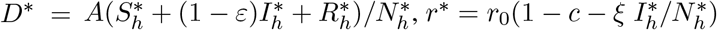 and 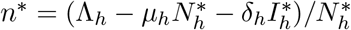, we have

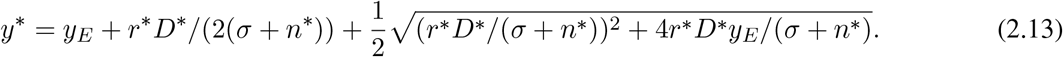

Therefore, any equilibrium value of *y* (*y*^*\**^) satisfies Eq. (2.13). The set of *y*^*\**^ values, and consequently all endemic equilibria of the model (2.1), are governed by solutions to Eq.(2.13). Since Eq. (2.13) can not be solved explicitly to obtain closed form solutions, the stability of endemic equilibria to the model (2.1) will be investigated numerically.

## 3. Numerical results

In this section, the model (2.1) is simulated to gain insights into the interplay between malaria dynamics and economic growth, assess the impact of some critical model parameters (including parameters through which some control measures can be evaluated) on key response functions such as the basic reproduction number, the infectious human population, and the per capita capital, as well as evaluating the possibility of long transient events, with a specific focus on long transients, on both malaria dynamics and economic growth. Unless explicitly specified, the simulations are carried out using the baseline parameter values outlined in Table S1 in Section S2 of the SI.

### 3.1 Long term dynamics of the model system

#### 3.1.1 Threshold dynamics and backward bifurcation

The model system (2.1) is simulated using the baseline parameter values tabulated in Table S1 to illustrate the fact that the basic reproduction number (*R*_0_) is indeed a threshold value. Results of the simulations presented in Fig. 2 depict a locally asymptotically stable disease-free equilibrium (*DFE*) when *β*_*v*0_ = 0.2345 and *R*_0_ = 0.9977 (dotted curves in Fig. 2 (a)-(c)) and a stable endemic equilibrium (*EE*) when *β*_*v*0_ = 0.24 and *R*_0_ = 1.0211 (solid curves in Fig. 2 (a)-(c)). This confirms the fact that *R*_0_ is a threshold value. Thus, the disease free equilibrium is asymptotically stable if *R*_0_ *<* 1, while the disease approaches a positive endemic equilibrium when *R*_0_ *>* 1.

**Figure 2:**
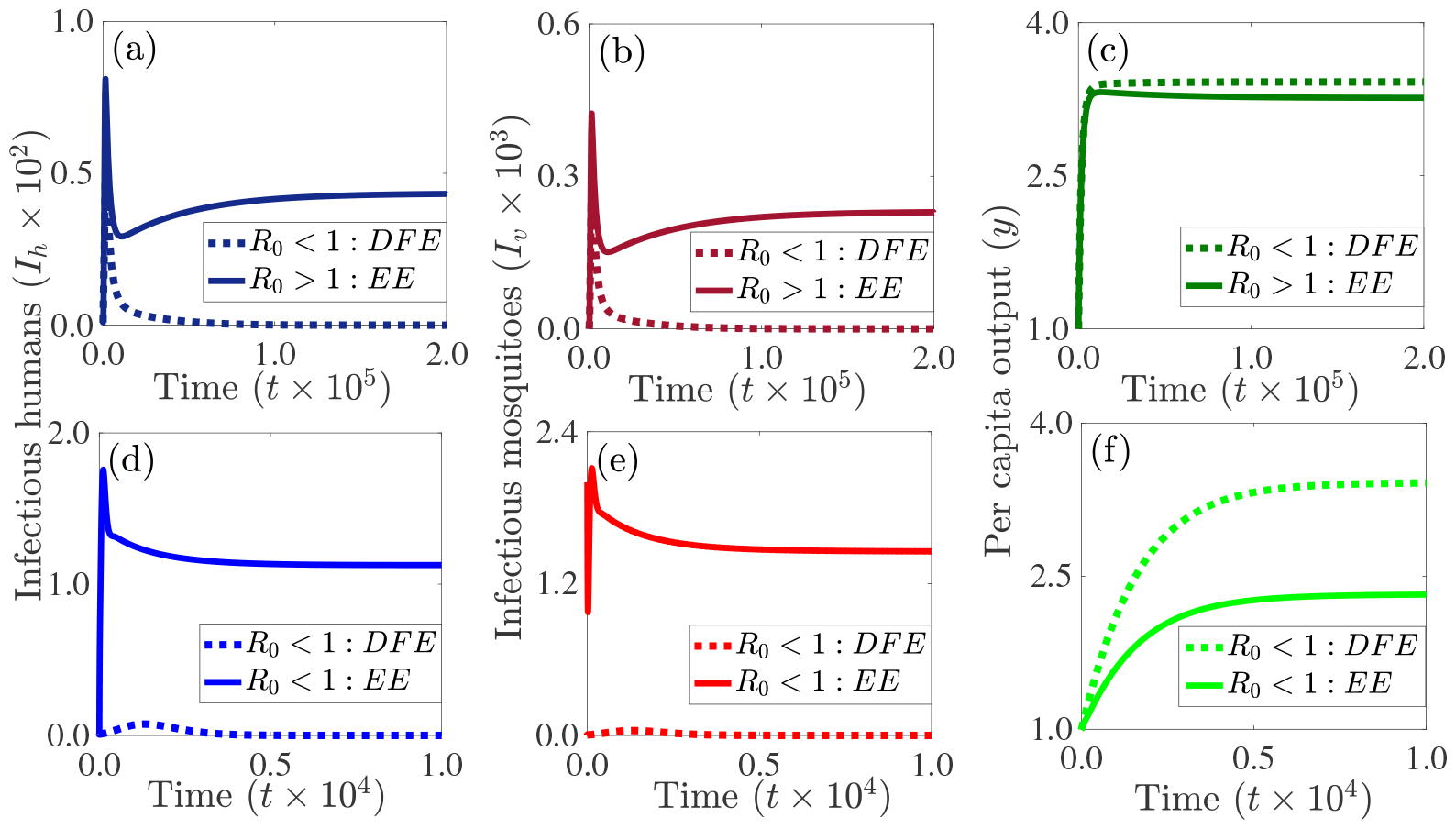
Simulations of the model (2.1) to confirm the fact that the basic reproduction number (*R*_0_) is a threshold value ((a)-(c)) and the existence of a backward bifurcation when *R*_0_ *<* 1 ((d)-(f)). When *β*_*v*0_ = 0.2345, the corresponding value of the basic reproduction number is *R*_0_ = 0.9977 *<* 1 and the disease-free equilibrium (*DFE*) of the model is locally asymptotically stable (dotted curves in (a)-(c)). When *β*_*v*0_ = 0.24, the corresponding value of the basic reproduction number is *R*_0_ = 1.0211 *>* 1 and the endemic equilibrium (*EE*) of the model is locally asymptotically stable (solid curves in (a)-(c)). The initial condition used for the simulations is (*S*_*h*_(0), *I*_*h*_(0), *R*_*h*_(0), *S*_*v*_(0), *I*_*v*_(0), *k*(0)) = (1499, 1, 0, 10000, 0, 1). For (d)(f), when *R*_0_ *<* 1, there is a parameter regime within which trajectories originating within the basin of attraction of the *DFE*, e.g., (*S*_*h*_(0), *I*_*h*_(0), *R*_*h*_(0), *S*_*v*_(0), *I*_*v*_(0), *k*(0)) = (1490, 1, 0, 10000, 0, 1), converge to the *DFE* (dotted curves in (d)-(f)), while trajectories originating within the basin of attraction of the stable *EE*, e.g., (*S*_*h*_(0), *I*_*h*_(0), *R*_*h*_(0), *S*_*v*_(0), *I*_*v*_(0), *k*(0)) = (490, 1, 0, 5000, 2000, 1), converge to the stable *EE* (solid curves in (d)-(f)). That is, a stable *DFE* and a stable *EE* co-exist for a parameter regime in which *R*_0_ *<* 1.

The values of the mosquito biting rate and human disease-induced death rates are 0.22 and 4.5068*×* 10^*−*4^, respectively, and the corresponding value of the reproduction number is 0.9278. The values of the other parameters are presented in Table S1.

Additionally, the model (2.1) is simulated using the baseline parameter values in Table S1, with a background mosquito biting rate *β*_*v*0_ = 0.22 and human disease-induced death rate *δ*_*h*_ = 4.5068*×* 10^*−*4^ *>* 4.9813*×* 10^*−*5^ = *µ*_*h*_ (the human natural death rate) to demonstrate the possibility of a backward (sub-critical) bifurcation. The results of the simulations depicted in Fig. 2 suggest the potential for the model (2.1) to exhibit a backward bifurcation, wherein a stable disease-free equilibrium (dotted curves in Fig. 2 (d)-(f)) coexists with a stable endemic equilibrium (solid curves in Fig. 2 (d)-(f)) within the same parameter regime for which the reproduction number is less than one. The occurrence of this phenomenon in the model (2.1) hinges on a substantial disparity between the disease-induced mortality rate and the natural mortality rate. While formal proof is not provided here, the Center manifold theory offers a rigorous approach for establishing the existence of a backward bifurcation [68]. It should be noted that the existence of a backward bifurcation signifies that, although the requirement that the reproduction number be less than one to contain a disease is necessary, it is not sufficient for achieving disease elimination (defined as a significant reduction in malaria cases to minimal or near-zero levels). In models with backward bifurcations, mere reduction of the reproduction number slightly below one, as is the case in models without backward bifurcations, does not guarantee disease elimination. Instead, in models with backward bifurcations, successful disease elimination necessitates intensive and sustained control and mitigation efforts until the reproduction number falls below a critical threshold value. That is, disease elimination occurs within the region where the disease-free equilibrium becomes globally asymptotically stable. In this backward bifurcation (i.e., bistability) scenario, the economic output associated with the endemic equilibrium is notably low, whereas the output linked to the disease-free equilibrium is relatively high. This demonstrates the negative impact of the disease on economic output, as well as the positive impact of a strong economy on disease control and mitigation.

#### 3.12 The impact of some key parameters on the long term dynamics of the model

The model (2.1) is simulated using the baseline parameter values presented in Table S1 to assess the impact of technological progress or labor efficiency (*A*) on the long term dynamics of the infectious human population (*I*_*h*_), the infectious mosquito population (*I*_*v*_), and the per capita yield (*y*). Results of the simulations show that increasing labor efficiency from its baseline value of 1 to 10 will lead to a significant reduction in the basic reproduction number from *R*_0_ *≈*4.04 to *R*_0_ *≈*0.97 *<* 1 and a significant increase in the equilibrium value of per capita yield (comparing the blue and green curves in Fig. 3 (a), (f), and (k)). However, reducing labor efficiency from its baseline value by 50% will lead to an *≈*6% increase in the endemic equilibrium value of infectious human population (comparing the blue and red curves in Fig. 3 (a)), an *≈*13% increase in the endemic equilibrium value of the infectious mosquito population (comparing the blue and red curves in Fig. 3 (f)), and an *≈*22% decrease in the equilibrium value of the per capita yield (comparing the blue and red curves in Fig. 3 (k)). For this scenario, there is a 22% increase in the basic reproduction number and the disease is endemic since (*R*_0_ *>* 1). Furthermore, increasing labor efficiency from its baseline value by 50% will lead to an *≈*5% decrease in the endemic equilibrium value of infectious human population (comparing the blue and gold curves in Fig. 3 (a)), an *≈*11% decrease in the endemic equilibrium value of the infectious mosquito population (comparing the blue and gold curves in Fig. 3 (f)), and an *≈*21% increase in the equilibrium value of the per capita yield (comparing the blue and red curves in Fig. 3 (k)). For this scenario, there is a 8% reduction in the basic reproduction number.

**Figure 3:**
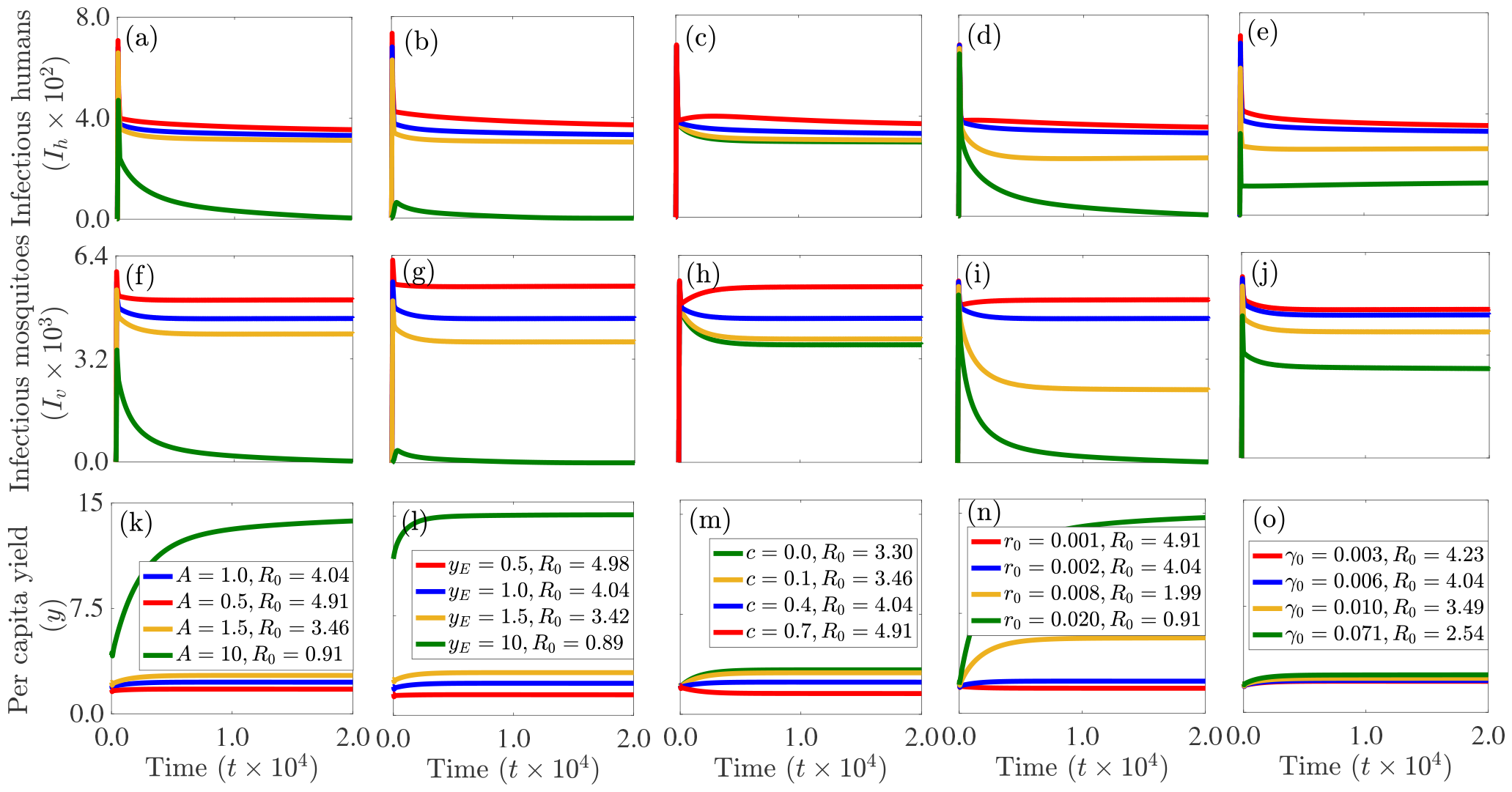
Simulations of the model (2.1) depicting the long term dynamics of the (a)-(e): infectious human population (*I*_*h*_), (f)-(j): infectious mosquito population (*I*_*v*_), and (k)-(o): per capita yield (*y*) for different values of technological progress or labor efficiencies (*A*: (a), (f), and (k)), external aid (*y*_*E*_: (b), (g), and (l)), proportion of yield consumed (*c*: (c), (h), and (m)), background investment rate (*r*_0_: (d), (i), and (n)), and background recovery rate (*γ*_0_: (e), (j), and (o)). The initial condition used for the simulations is (*S*_*h*_(0), *I*_*h*_(0), *R*_*h*_(0), *S*_*v*_(0), *I*_*v*_(0), *k*(0)) = (999, 1, 0, 10000, 0, 1), while the values of the other parameters are presented in Table S1.

Also, the model (2.1) is simulated to assess the impact of external aid (*y*_*E*_) on the long term dynamics of the system. The results obtained and illustrated in Fig. 3 (b), (g), and (l) show that for the baseline parameter values in Table S1, the reproduction number is *R*_0_ *≈* 4.04 and the corresponding dynamics are illustrated by the blue curves in Fig. 3. Reducing external aid from its baseline value of 1.0 by 50% leads to an *≈*23% increase in the reproduction number, an 11% increase in the endemic equilibrium value of the infectious human population, an *≈*22% increase in the endemic equilibrium value of the infectious mosquito population, and an *≈*36% reduction in the equilibrium value of per capita yield (comparing the blue and green curves in Fig. 3 (b), (g), and (l)). However, increasing external aid by 50% will trigger an *≈*15% decrease in the reproduction number, an 8% decrease in the endemic equilibrium value of the infectious human population, an *≈*16% decrease in the endemic equilibrium value of the infectious mosquito population, and an *≈*33% increase in the equilibrium value of per capita yield (comparing the blue and gold curves in Fig. 3 (b), (g), and (l)). A significant increase in external aid can result in disease containment. Specifically, increasing external aid from its baseline value to 1 to 10 will lead to a reproduction number of *R*_0_ *≈*0.89, i.e. an *≈*78% reduction in the baseline value of the reproduction number (comparing the blue and green curves in Fig. 3 (b), (g), and (l)).

Furthermore, the model (2.1) is simulated using the baseline parameter values presented in Table S1 to assess the impact of the portion of yield consumed (*c*) on the long term dynamics of the system. The results obtained and depicted in Fig. 3 (c), (h), and (m) show that for the baseline value of *c*, the model relaxes at an endemic equilibrium (blue curves in Fig. 3 (c), (h), and (m)). Increasing the baseline portion of the yield consumed by 75% will result in an *≈* 22% increase in the basic reproduction number, an *≈*10% ( *≈*22%) increase in the equilibrium infectious human (mosquito) population, and a 35% reduction in the per capita yield (comparing the blue and red curves in Fig. 3 (c), (h), and (m)). However, a 75% reduction in the baseline portion of the yield consumed will lead to an *≈*14% reduction in the basic reproduction number an *≈*7% (*≈*14%) reduction in the equilibrium infectious human (mosquito) population, and an *≈*29% increase in the per capita yield (comparing the blue and gold curves in Fig. 3 (c), (h), and (m)). Additional reductions in the proportion of the yield consumed will result in further reductions in the reproduction number, as well as the endemic infectious human and mosquito populations. However, it should be noted that this will not lead to disease containment, even when none of the yield is consumed. In particular, if none of the yield is consumed (i.e., if *c* = 0.0), an *≈*18% reduction in the basic reproduction number, an *≈*9% ( *≈*19%) increase in the equilibrium infectious human (mosquito) population, and an *≈*39% reduction in the per capita yield (comparing the blue and green curves in Fig. 3 (c), (h), and (m)).

Additional simulations of the model (2.1) are carried out using the baseline parameter values presented in Table S1 to assess the impact of the background investment rate in capital (*r*_0_) on the long term dynamics of the system. The results obtained and depicted in Fig. 3 (d), (i), and (n) show that halving the baseline background investment rate in capital will trigger an *≈*22% increase in the basic reproduction number, an *≈*6% ( *≈*13%) increase in the equilibrium infectious human (mosquito) population, and a 22% reduction in the per capita yield (comparing the blue and red curves in Fig. 3 (d), (i), and (n)), while significant increases in the baseline background investment rate in capital will generate significant reductions in the infectious human and mosquito equilibria and significant increases in the per capita yield. Specifically, increasing the baseline background investment rate in capital ten-fold will lead to a reproduction number that is less than unity and the system will converge to the disease-free equilibrium (comparing the blue and green curves in Fig. 3 (d), (i), and (n)). In summary, notable increases in the baseline background investment rate in capital result in significant reductions in the infectious human and mosquito equilibria. In practical terms, this implies that higher investments in economic capital, including improved infrastructure, drainage systems, sanitation, healthcare facilities, or economic development projects, contribute to reduced disease transmission levels. For example, improved economic conditions may enable individuals and communities to implement effective mosquito control and mitigation measures, invest in healthcare infrastructure, and implement public health interventions, leading to reduced disease prevalence and transmission.

Fig. 3 (e), (j), and (o) depicts simulations of the model (2.1) using the baseline parameter values given in Table S1 to assess the impact of the human background recovery rate in from infection (*γ*_0_) on the long term dynamics of the system. They indicate that reducing the baseline value of the human background recovery rate in from infection by half will lead to an *≈* 5% increase in the basic reproduction number, an *≈* 6% (*≈* 4%) increase in infectious human (mosquito) endemic equilibrium, an (*≈*3%) reduction in per capita yield (comparing the blue and red curves in Fig. 3 (e), (j), and (o)). On the other hand, increasing the human background recovery rate in from infection will result in decreases in basic reproduction number and the equilibrium infectious human and mosquito populations. For example, increasing the baseline value of *γ*_0_ to 0.0714 (i.e., setting the average duration of infection to 14 days) will result in an *≈* 37% reduction in the basic reproduction number, an *≈*59% ( *≈*38%) reduction in the infectious human (mosquito) endemic equilibrium, and an *≈*18% increase in per capita yield (comparing the blue and red curves in Fig. 3 (e), (j), and (o)).

More simulations were carried out to assess the impacts of the malaria-related medical costs parameter (*ξ*) and the adjustment factor for decreased productivity associated with clinical malaria (*ε*) on the long term dynamics of the model (2.1). The results obtained and presented in Fig. S1 of the SI show that more expenditure on malaria will lead to reduced per capita yield and increased malaria prevalence, while reduced spending on malaria will lead to increased per capita yield and reduced malaria prevalence. In particular, a 50% increase in the baseline value of *ξ* will generate an *≈* 7.1% reduction in per capita yield and an *≈*1.9% ( *≈*4.1%) increase in the infectious human (mosquito) equilibrium (comparing the blue and red curves in Fig. S1 (a)-(c)), while reducing the baseline spending on malaria by half will lead to an *≈*6.5% increase in per capita yield and an *≈*1.7% ( *≈*3.6%) reduction in the infectious human (mosquito) equilibrium (comparing the blue and gold curves in Fig. S1 (a)-(c)). More increases in per capita yield ( *≈* 12.5%) and more reductions in malaria prevalence (*≈*3.6% for humans and *≈*6.6% for mosquitoes) are achieved if no portion of the yield is spent on malaria-related illness (comparing the blue and green curves in Fig. S1 (a)-(c)). Similarly, a higher decrease in economic productivity caused by malaria will lead to a decrease in per capita yield and an increase in malaria prevalence, whereas a lower decrease in economic productivity due to malaria will result in an increase in per capita yield and a decrease in malaria prevalence. Specifically, if no clinically sick human contributes to productivity (i.e., if *ε* = 1), an *≈*9.9% reduction in per capita yield and an *≈*2.7% ( *≈*5.6%) increase in the infectious human (mosquito) equilibrium is recorded (comparing the blue and red curves in Fig. S1 (d)-(f)), while if everybody including clinically sick humans contribute to productivity (i.e., if *ε* = 0) an *≈*8.9% increase in per capita yield and an *≈*2.3% (*≈* 4.7%) reduction in the infectious human (mosquito) equilibrium is recorded (comparing the blue and green curves in Fig. S1 (d)-(f)).

### 3.2 Assessing the impact of key parameters on the basic reproduction number

In this section, heat maps are generated to assess the influence of critical parameters of the model (2.1) on disease control, quantified by the basic reproduction number (*R*_0_) using the baseline parameter values from Table S1 (unless otherwise specified). The outcomes are illustrated in Fig. 4.

**Figure 4:**
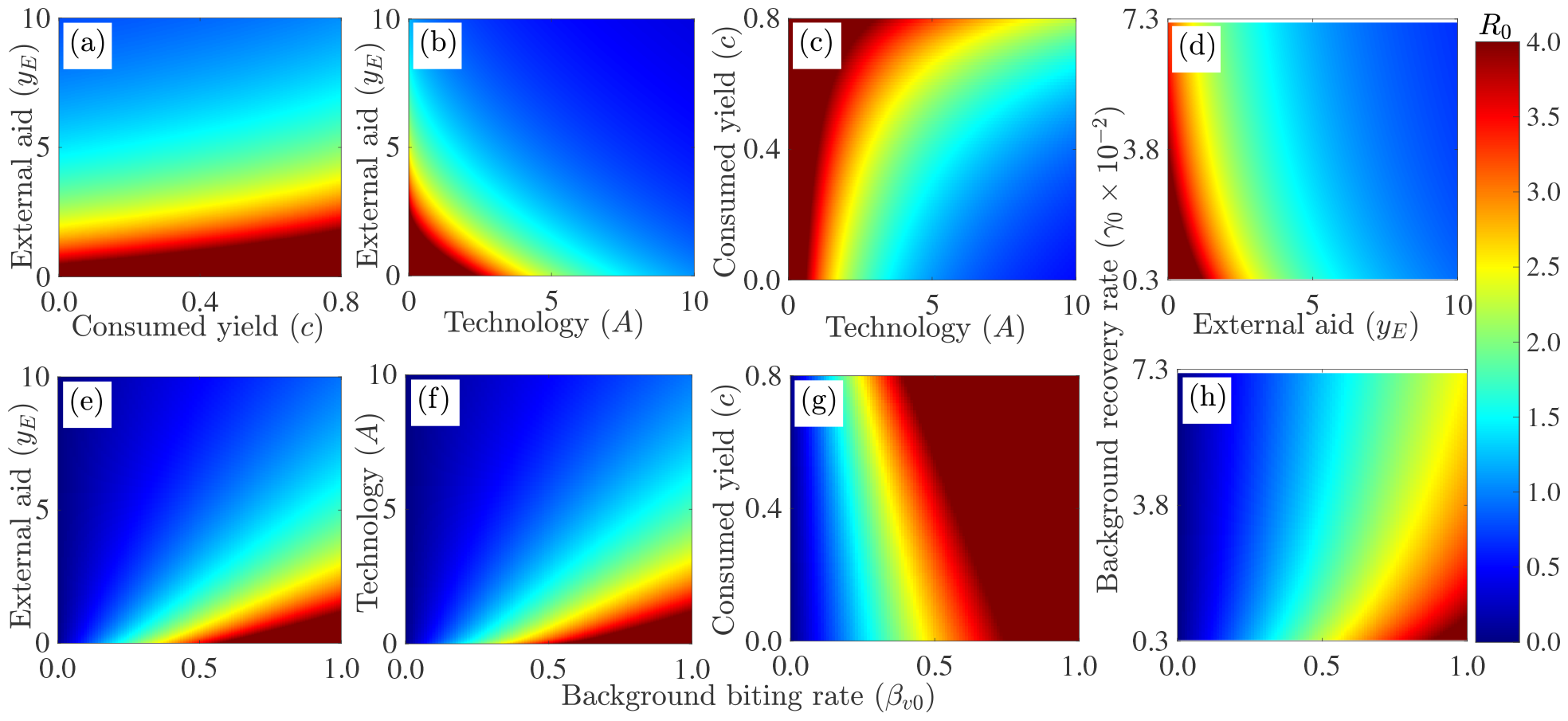
Heatmaps of the basic reproduction number (*R*_0_) of the model (2.1) as a function of (a) the fraction of the yield consumed (*c*) and external aid (*y*_*E*_), (b) technological progress or labor efficiency (*A*) and external aid (*y*_*E*_), (c) technological progress and the fraction of the yield consumed, (d) external aid and background recovery rate (*γ*_0_), (e) background mosquito biting rate (*β*_*v*0_) and external aid, (f) background mosquito biting rate and technological progress, (g) background mosquito biting rate and the fraction of the yield consumed, and (h) background mosquito biting rate and the background recovery rate. With the exception of these varied parameters, the other parameters are held at their baseline values given in Table S1.

Figure 4 (a) depicts a heat map of the basic reproduction number (*R*_0_) as a function of the fraction of the yield that is consumed (*c*) and external aid (*y*_*E*_). The heat map indicates that achieving a reproduction number that is below one (which is required to control the disease) is intricately linked to both the level of external aid and the proportion of the yield consumed. Specifically, if both consumption and external aid are maintained at their baseline values, it becomes unfeasible to reduce the reproduction number below one. In contrast, if none of the yield is consumed, achieving a reproduction number that is below one necessitates less external assistance compared to when a portion of the yield is consumed. In particular, upholding the consumed portion of the yield at its baseline value requires an extra 15% augmentation in the external aid value, linked to zero consumed yield, to attain a reproduction number reduction below one. Moreover, elevating the baseline fraction of the yield consumed by 50% demands an additional 24% increase in the external aid value, associated with zero consumed yield, to achieve a reduction in the reproduction number below one. In summary, a higher consumption of yield complicates the task of reducing the reproduction number below one. When consumption is exceptionally high and external aid is minimal, achieving *R*_0_ *<* 1 becomes unattainable. These findings underscore the intricate interplay between external aid, yield consumption, and their collective impact on the reproduction number (a crucial determinant in infectious disease dynamics), providing valuable insights into the dynamics of disease transmission and prevention.

Figure 4 (b) presents a heat map illustrating the relationship between the basic reproduction number (*R*_0_), technological progress (*A*), and external aid (*y*_*E*_). This plot reveals that when reducing the basic reproduction number below one is impossible if both technological progress and external aid are maintained at their baseline values. However, if technological progress is held at its baseline value, a 8.8-fold increase in the baseline value of external is required to reduce the basic reproduction number below one, whereas if external aid is held at its baseline value, a 9-fold increase in technological progress is required to reduce the basic reproduction number below one. On the other hand, a four-fold increase in baseline value of external aid requires a five-fold increase in baseline value of technological progress to reduce the reproduction number below one. Hence, if external aid is high, less external aid is needed to bring the basic reproduction number below one. Conversely, with higher external aid, less technological progress is required to achieve this goal. In summary, higher technological progress reduces the need for external aid in controlling the disease; similarly, increased aid lessens the required technological progress.

Additionally, a heat map of the basic reproduction number as a function of technology and the fraction of the yield that is consumed (Figure 4 (c)) is used assess the combined impact of these parameters on disease control. The results obtained show that reducing the reproduction number below one is unachievable if both parameters are held at their baseline values stipulated in Table S1. However, if no portion of the yield is consumed, a threshold level of technology (which is approximately 5.5 times the baseline value of technological progress) is identified, allowing the reproduction number to fall below one. If the portion of the yield consumed is maintained at its baseline value, a nine-fold increase in the baseline value of technological progress is required to achieve a reduction in the reproduction number below one. Alternatively, if the baseline value of the consumed yield is reduced by 50%, a seven-fold increase in the baseline value of technological progress is needed to attain the same outcome. To summarize, reducing the reproduction number below one is challenging at baseline values, but achievable with specific adjustments in technology and yield consumption.

Figure 4 (d) shows a heat map of the basic reproduction number as a function of external aid and the human background recovery rate from infection. The graphical representation illustrates that achieving disease containment is unattainable when both external aid and the background recovery rate are kept at their baseline values, as outlined in Table S1. But containing the disease might be feasible if the background recovery rate is maintained at its baseline value but external aid is increased. In summary, disease containment is unattainable when maintaining both external aid and the background recovery rate at baseline values, but there is a potential for containment by increasing external aid while keeping the background recovery rate constant.

Figures 4 (e)-(h) illustrate heat maps representing the basic reproduction number (*R*_0_) of model (2.1). These plots demonstrate its relationship with the background biting rate of mosquitoes (*β*_*v*0_) and external aid (Fig. 4 (e)), technological progress (Fig. 4 (f)), the fraction of the yield consumed (Fig. 4 (g)), and the background recovery rate (Fig. 4 (h)). The findings highlight the challenges associated with malaria control in regions characterized by high mosquito biting rates, particularly in the presence of limited external aid, low technological advancement, inadequate treatment, or high consumption of the yield. Specifically, if the background biting rate of mosquitoes is held at its baseline value in Table S1, high external aid or technology is required to reduce the reproduction number below one, while it becomes impractical to reduce the reproduction number below one and thus contain the disease effectively, even when none of the yield is consumed or for a very high background recovery rate.

It should be noted that reducing the mosquito biting rate, along with increasing external aid, technology, or the background recovery rate, has a positive impact on lowering the reproduction number. In the absence of external aid, achieving a basic reproduction number below one requires a reduction of approximately 85% in the baseline background biting rate of mosquitoes, with smaller reductions needed in the presence of external aid (Fig. 4 (e)). Specifically, with baseline external aid, a 75% reduction is necessary, and a fourfold increase in external aid requires only a 47% reduction in the baseline mosquito biting rate for the same outcome (Fig. 4 (e)). Additionally, increasing external aid fourfold leads to a basic reproduction number below one, even with the baseline mosquito biting rate (Fig. 4 (e)). Similar trends are observed for technological progress (Fig. 4 (f)). Furthermore, simultaneously reducing both the mosquito biting rate and the fraction of the yield consumed accelerates disease control. Specifically, maintaining the consumed portion of the yield at its baseline value demands an 85% reduction in the baseline mosquito biting rate to achieve a basic reproduction number below one. If the consumed portion is increased by 75%, an 88% reduction in the mosquito biting rate is required (Fig. 4 (f)). If none of the yield is consumed, an 81% reduction in the mosquito biting rate is necessary (Fig. 4 (f)). Figure 4 (g) demonstrates that holding the background recovery rate of humans at its baseline value requires a 76% reduction in the mosquito biting rate for a basic reproduction number below one. Increasing the baseline human recovery rate to *≈*0.0714 (i.e., recovery in two weeks) necessitates a 59% reduction in the mosquito biting rate to achieve the same outcome. In conclusion, controlling malaria in areas with high mosquito biting rates is challenging, especially when facing limited external aid, low technology, inadequate treatment, or high yield consumption. Reducing mosquito biting plays a crucial role in disease control, especially when coupled with external aid and advanced technology. It should be mentioned that, reducing mosquito biting can be achieved through vector control measures like Long-lasting insecticide-treated nets, while increased recovery rates can result from enhanced diagnosis and treatment.

### 3.3 External aid distribution strategy

The model (2.1) is simulated to assess the impact of various external aid distribution strategies on the infectious human and mosquito populations, as well as on per capita yield. The strategies involve the distribution of a fixed amount of external aid over three-year periods. **Strategy 1**: Full allocation exclusively in the first year, with no external aid in the subsequent two years (Fig. 5 (a)-(c)). **Strategy 2**: Equitable annual distributions over the first two years with no allocation in the third year (Fig. 5 (d)-(f)). **Strategy 3**: Equal annual allocations across the three years (Fig. 5 (g)-(i)). Each of the strategies is repeated for three years over a 15-year period. These strategies are assessed in conjunction with a scenario where no external aid is allocated (magenta curves in Fig. 5).

**Figure 5:**
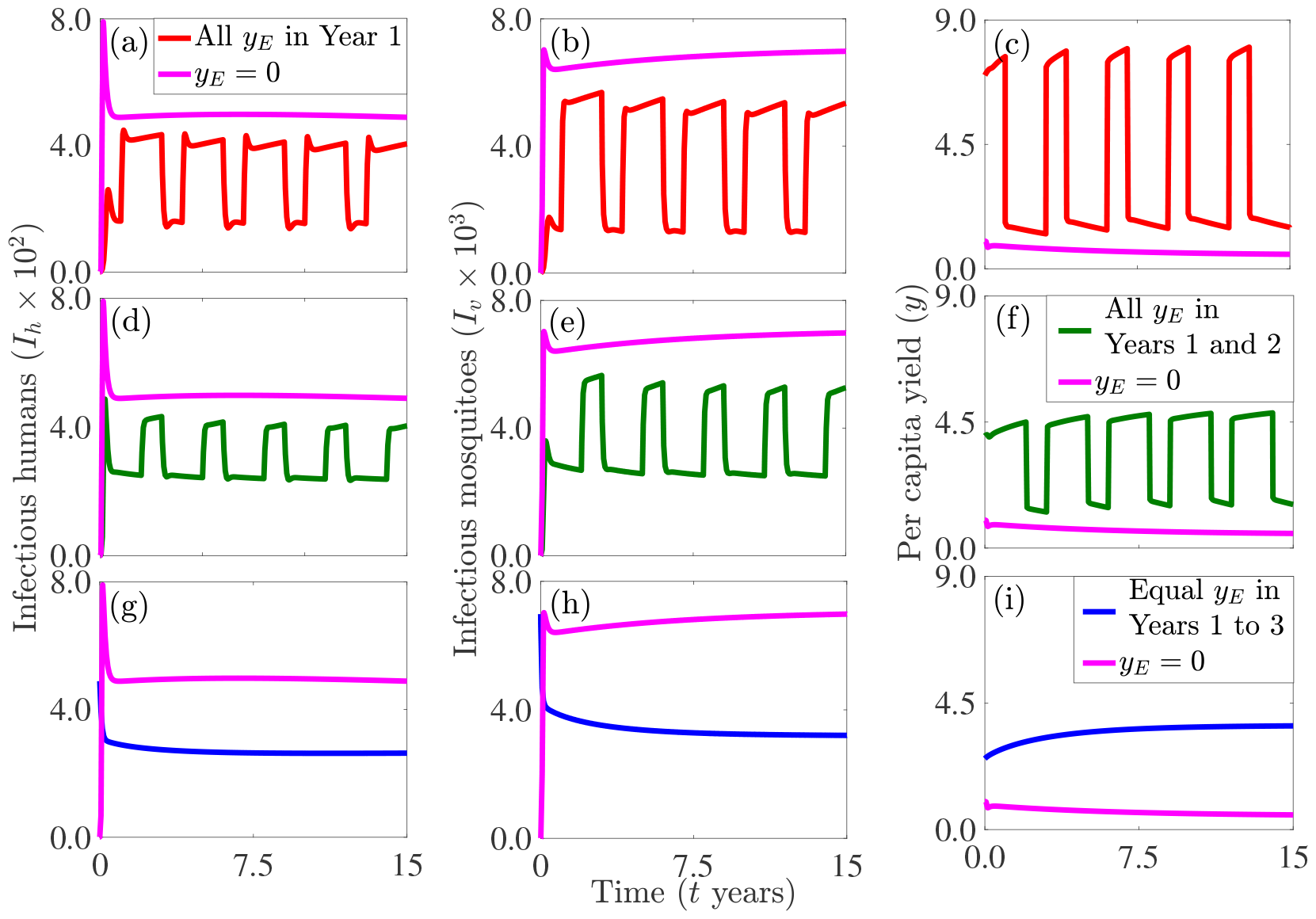
Simulations of the model (2.1) to assess various external aid (*y*_*E*_) allocation strategies. (a)-(c): All external aid is distributed in the first year (with no external aid in Years 2 to 3). (d)-(f): All external aid is distributed evenly on an annual basis over the first two years (with no external aid in Year 3). (d)-(f): All external aid is distributed evenly on an annual basis over three years. The initial condition used for the simulations is (*S*_*h*_(0), *I*_*h*_(0), *R*_*h*_(0), *S*_*v*_(0), *I*_*v*_(0), *k*(0)) = (999, 1, 0, 10000, 0, 1), while the values of the other parameters are outlined in Table S1.

Strategy 1 compared with the no external aid scenario results in a 35% (42%) reduction in the total infectious human (mosquito) population and a 470% increase in the total per capita yield over the 15-year period (comparing the areas under the magenta and red curves in Fig. 5 (a)-(c)). Similarly, for Strategy 2, a 40% (48%) reduction in the total infectious human (mosquito) population and a 462% increase in the total per capita yield is recorded over the three-year period (comparing the areas under magenta and green curves in Fig. 5 (d)-(f)). For Strategy 3, a 46% (50%) reduction in the total infectious human (mosquito) population and a 445% increase in the total per capita yield is observed over the three-year period (comparing the areas under magenta and blue curves in Fig. 5 (g)-(i)). Comparing the results of the various strategies, implementing Strategy 2 instead of Strategy 1 results in an additional 5% (6%) reduction in the total infectious human (mosquito) population and a 8% reduction in the total per capita yield over the 15-year period (comparing the areas under red and green curves in Fig. 5 (a)-(c) and Fig. 5 (d)-(f)). Implementing Strategy 3 instead of 1 results in an additional 11% (8%) reduction in the total infectious human (mosquito) population and an additional 25% decrease in the total per capita yield over the three-year period (comparing the areas under red and green curves in Fig. 5 (a)-(c) and Fig. 5 (g)-(i)), while implementing Strategy 3 instead of 2 results in an additional 6% (2%) reduction in the infectious human (mosquito) population and a 17% decrease in the total per capita yield over the three-year period (comparing the areas under gold and green curves in Fig. 5 (d)-(f) and Fig. 5 (g)-(i)). In summary, although each of the three strategies results in a decreased total number of cases and an increased per capita yield, Strategy 3 involving equitable annual allocation of external aid leads to the lowest total number of disease cases and the lowest total per capita yield over the 15-year period.

Additional simulations were carried out to assess the impact of diverse external aid allocation strategies, considering different time frames and durations, on the total infectious human population over a 15-year period, with each strategy repeated every three years. Given the overlap between previous larger distribution time points and some of the smaller distribution time points in subsequent strategies, we avoid repeating them, except for the full equitable distribution points in this discussion. The distribution scenarios included: 1) Yearly distributions–all external aid distributed during the first year with no external aid during the second and third years (blue dots in Fig. 6), two equal installments during the first two years and no external aid during the third year (blue squares in Fig. 6), and three equal installments (blue diamonds in Fig. 6). 2) Bi-annual distributions–bi-annually but only during the first six months (magenta dots in Fig. 6), bi-annually but only during the first 18 months or 1.5 years (magenta squares in Fig. 6), and six equal installments (blue diamonds in Fig. 6). 3) Quarterly distributions–quarterly but only during the first quarter (green dots in Fig. 6), quarterly but only during the first 3 quarters (green pentagrams in Fig. 6), quarterly but only during the first 5 quarters (green hexagrams in Fig. 6), quarterly but only during the first 7 quarters (green squares), quarterly but only during the first 9 quarters (green triangles in Fig. 6), quarterly but only during the first 11 quarters (green diamonds in Fig. 6), and twelve equal quarterly installments (blue diamonds in Fig. 6). 4) Monthly distributions–all external aid distributed only during the first month (orange dot in Fig. 6 (d)), first two months, first three months, etc., and upto 36 equal monthly installments (blue diamond in Fig. 6 (d)).

**Figure 6:**
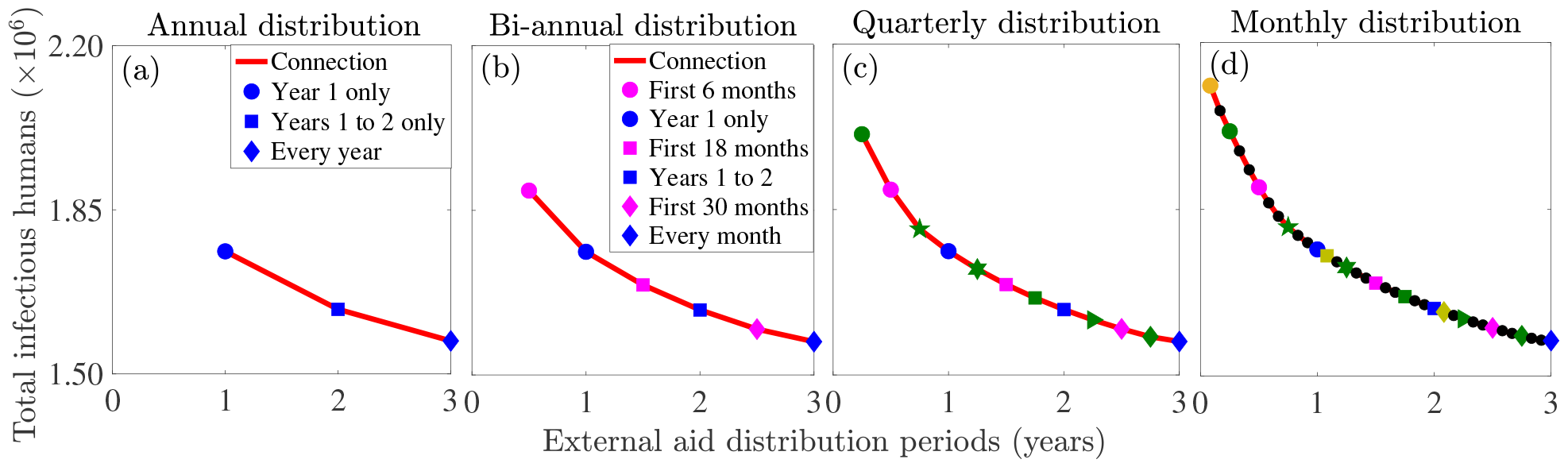
Simulations of the model (2.1) to assess various external aid (*y*_*E*_) strategies. The initial condition used for the simulations is (*S*_*h*_(0),*I*_*h*_(0),*R*_*h*_(0),*S*_*v*_(0),*I*_*v*_(0),*k*(0)) = ( 999,1,0,10000,0, 1), while the values of the other parameters are outlined in Table S1.

The findings reveal that an effective strategy for optimized disease control entails distributing available aid evenly. In particular, allocating all external aid within the initial six months results in a 7% higher total infectious human population compared to distributing all external aid solely in the first year (comparing the magenta dot in Fig. 6 (b) with the blue dot in Fig. 6 (a)). Similarly, allocating all external aid within the first quarter leads to a 14% higher total infectious human population compared to distributing all external aid solely in the first year (comparing the green dot in Fig. 6 (c) with the blue dot in Fig. 6 (a)). Additionally, allocating all external aid within the first month results in a 20% higher total infectious human population compared to distributing all external aid solely in the first year (comparing the orange dots in Fig. 6 (d) with the blue dot in Fig. 6 (a)). Nonetheless, if all external aid is distributed in equal annual installments for two years only, an 8% reduction in the total infectious human population compared to allocating all external aid solely during the first year will be recorded (comparing the blue dot and square in Fig. 6 (a)), while if all external aid is distributed in four six-month installments only, a 16% reduction in the total infectious human population compared to allocating all external aid solely during the initial six months will be recorded (comparing the magenta dot and blue square in Fig. 6 (b)). Similarly, if all external aid is distributed in eight quarterly installments, a 23% reduction in the total infectious human population compared to allocating all external aid during the first quarter will be recorded (comparing the green dot and blue square in Fig. 6 (c)). Furthermore, distributing all external aid in 24 monthly installments will result in a 29% reduction in the total infectious human population compared to distributing all external aid solely during the first month (comparing the orange dot and blue square in Fig. 6 (d)). Similar reductions are observed as the distribution frequency increases. Precisely, distributing all aid in equal annual installments leads to an 11% reduction in the total number of infectious humans compared to distributing all aid during the first year only (comparing the blue dot and diamond in Fig. 6 (a)). Distributing all aid equally every six months results in a 17% reduction in the total number of infectious humans compared to distributing all aid during the first six months only (comparing the magenta dot and blue diamond in Fig. 6 (b)). Distributing all aid in equal quarterly installments leads to a 22% reduction in the total number of infectious humans compared to distributing all aid during the first quarter only (comparing the green dot and blue diamond in Fig. 6 (c)), while distributing all aid in equal monthly installments results in a 26% reduction in the total number of infectious humans compared to distributing all aid during the first month only (comparing the orange dot and blue diamond in Fig. 6 (d)). In summary, allocating all external aid within six, three, or one month leads to a higher total number of infectious cases compared to distributing external aid within one year, while equitable distribution of external aid leads to the lowest total number of infectious humans. The most substantial reduction in total infectious cases is observed with the equal monthly distribution strategy compared to equal annual, bi-annual, and quarterly distribution strategies.

### 3.4 Long transient dynamics for the coupled system

In mathematics and ecology, “long transients” refer to extended periods during which a dynamic system, such as a mathematical model or an ecological population, takes a substantial amount of time to reach a stable or equilibrium state. These transients occur due to complex interactions, feedback loops, or time delays within the system, preventing it from quickly reaching a steady state. Long transients are often observed in ecological models where populations respond to changing environmental conditions or perturbations, and in mathematical systems exhibiting complex non-linear behavior before eventually converging to an attractor or a stable solution. Understanding long transients is essential for predicting the behavior of dynamic systems over time and for studying the factors that influence their transitions to stability or new dynamic regimes. In epidemiology, long transients can be relevant when studying the dynamics of infectious diseases within populations. They can help researchers understand how disease prevalence varies over extended periods before stabilizing or evolving into new patterns. This understanding is essential for effective disease control and intervention planning. In particular, in infectious disease modeling, long transients can signal the existence of reservoirs of infection. These are subpopulations or environmental factors that maintain the disease even during periods when it seems to be under control. Identifying and addressing such reservoirs is vital for preventing disease resurgence. Figure. 7 demonstrates the occurrence of this phenomenon in our model system when the initial conditions are near a saddle point. The system converges to the disease-free equilibrium (dotted curves in Fig. 7) and to the endemic equilibrium (solid curves in Fig. 7), with only a slight change in the initial susceptible human population. These results are attainable for the same parameter regime for which a backward bifurcation occurs. That is, for a high value of the disease-induced mortality rate (*δ*_*h*_) and a low value of the background mosquito biting rate (*β*_*v*0_). These long transient dynamics, characterized by the system staying near the saddle point for a prolonged time before transitioning to the endemic equilibrium is sensitive to the initial condition. That is, the region for the model (2.1) exhibits long transient dynamics is very small, suggesting that this phenomenon is rare for the malaria model. It should be emphasized that interpreting the system’s prolonged stay at the saddle point as a stable equilibrium can be misleading, potentially masking the impending transition to a high-level infection state.

**Figure 7:**
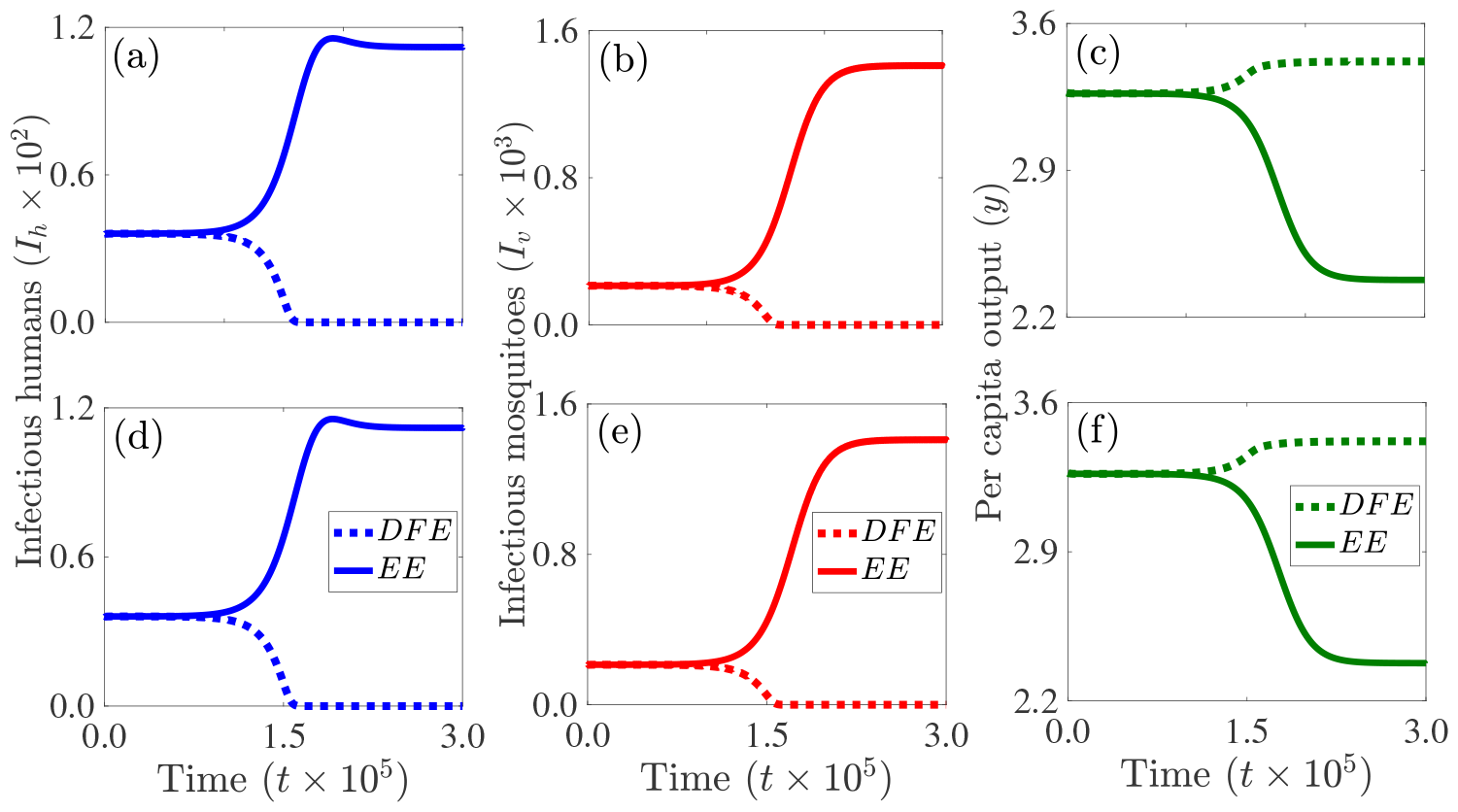
Simulations of the model (2.1) illustrating the long transients phenomena. The system converges to a disease-free equilibrium (*DFE*) denoted by dotted curves for the initial conditions: (*S*_*h*_(0), *I*_*h*_(0), *R*_*h*_(0), *S*_*v*_(0), *I*_*v*_(0)) = (1072.96, 36.08, 64.45, 6202.61, 213.17, 3.27) and to an endemic equilibrium (*EE*) denoted by solid curves for the initial conditions: (*S*_*h*_(0),*I*_*h*_(0),*R*_*h*_(0),*S*_*v*_(0),*I*_*v*_(0)) = (1072.94, 36.08, 64.45, 6202.61, 213.17, 3.27). For this simulation, *β*_*v*0_ = 0.22, *δ*_*h*_ = 4.5068 *×* 10^*−*4^, and the other parameters are maintained at their baseline values stipulated in Table S1. The reproduction number for this parameter regime is 0.936. It should be noted that the system stays near the saddle point for a prolonged time ((a)-(c)) before transitioning to the endemic equilibrium ((d)-(f)).

## 4. Discussion, limitations, and conclusion

Malaria imposes significant annual costs encompassing both direct prevention and care expenses and indirect costs such as lost productivity. Economic considerations involve prioritizing malaria control, choosing tailored prevention and treatment strategies, evaluating options, and optimizing resource allocation to ensure efficiency, highlighting the crucial role of economics in decision-making. In economic growth studies, the impact of improved labor quality on income and economic productivity has been explored extensively, with a focus on education rather than health. Understanding the relationship between health investments and increased labor productivity, particularly in agricultural-based developing countries is crucial. Evidence is needed to demonstrate how reducing sickness-related work absences can enhance overall efficiency and productivity. Conversely, understanding how the economy improves health outcomes is essential. Using malaria as a prototype disease, a framework integrating malaria dynamics, socio-economic factors, and transient events is formulated and analyzed. Unlike conventional malaria models, this framework integrates the malaria model with an economic growth model. The linkage occurs through malaria-related medical costs affecting the investment term, human population growth rate influencing the capital depreciation term, and reduced productivity due to malaria in the production function. Conversely, the economic model is connected to the malaria model through the mosquito biting rate in the forces of infection, human recovery rate from infection, mosquito recruitment term, and the mosquito mortality rate. This framework is used to analyze the synergistic feedback between malaria dynamics and economic growth, while also assessing the impact of different external aid allocation strategies and transient events on both malaria dynamics and economic growth.

Analysis of the integrated framework shows that the reproduction number is a function of both epidemiological and economic parameters implying that the framework can be used to assess the impact of socio-economic factors and disease parameters on disease control. Similar to many traditional malaria models, when the reproduction number is below unity, there exists a parameter range where disease containment is possible and another range where a backward bifurcation occurs. As in [46, 49, 50, 52], the backward bifurcation occurs when the human disease-induced mortality rate is significantly higher than the human natural mortality rate. This backward bifurcation phenomenon implies that achieving disease elimination requires intensified and sustained control measures until the reproduction number drops below a critical threshold and hence a push to lower transmission levels. In particular, a backward bifurcation and hence bistability in disease dynamics has profound implications for intervention outcomes and the emergence or re-emergence of diseases. Minor parameter changes can trigger significant fluctuations in incidence. Elimination states may be robust, but relaxing control efforts can trigger sudden transitions, and disease-free regions may experience abrupt shifts based on local conditions determining transmission intensity. In this backward bifurcation (bistability) scenario, the economic output linked to the disease-free equilibrium is comparatively high, while the economic output associated with the stable endemic equilibrium is significantly low. This underscores the detrimental effect of the disease on economic output.

Further analysis of the framework reveals that in the case where the elasticity coefficient is 0.5, the framework exhibits a single interior endemic equilibrium when the reproduction number exceeds one in the presence of external aid. However, in the absence of external aid, multiple interior endemic equilibria are possible, which is consistent with simulation results in [54, 55, 69, 70]. For this case in which the reproduction number is bigger than one, invasion establishes high prevalence, while lower transmission intensities trap imported cases into low prevalence equilibria. The existence of endemic equilibria when *R*_0_ *>* 1, a disease-free equilibria when *R*_0_ *<* 1, and the possibility of a backward bifurcation are confirmed through numerical simulations. These simulations show that various disease and economic parameters (including labor efficiency, external aid, fraction of the yield consumed, background investment rate, and background recovery rate from malaria) impact both disease and economic outcomes, although their effects vary in magnitude and scope. Additionally, the simulations show a reciprocal relationship between malaria and per capita yield. On one hand, the presence of malaria significantly diminishes per capita yield, suggesting that individuals in regions with a high malaria burden are more likely to experience lower economic productivity. Conversely, the level of per capita yield plays a crucial role in reducing the prevalence of malaria. Higher economic productivity often leads to improved living conditions and greater access to resources, which can contribute to lower malaria prevalence rates in affected areas. This highlights the intricate interplay between disease dynamics and socio-economic factors in endemic regions. This is consistent with findings from [71] depicting a bidirectional relationship between malaria burden and economic development, with economic progress contributing to a decrease in malaria burden. This connection is notably reflected in the strong correlation between malaria burden and indicators such as GDP per capita and total health expenditure per capita. These findings align with those in [55], underscoring the importance of universal healthcare access for fostering economic growth.

Heatmaps are used to assess the combined impact of parameters on the reproduction number. The results show that controlling malaria in high mosquito biting areas is challenging with limited aid, low technology, inadequate treatment, or high yield consumption, emphasizing the intricate interplay between these parameters and disease dynamics. However, reducing mosquito biting, increasing aid, technology, or the recovery rate impacts lowering the reproduction number positively. Hence, reducing mosquito biting coupled with increased external aid or technology, is crucial for disease control even in high mosquito biting areas. Reducing mosquito biting can be achieved through measures like insecticide-treated nets, while enhanced diagnosis and treatment increase recovery rates. Furthermore, the study shows that increased technological progress reduces external aid reliance, and that increased external aid reduces the required technological progress for disease control. Hence, the study identifies important parameters that can be calibrated using available or newly collected data for proper model validation.

Moreover, simulations of the model (2.1) were carried out to evaluate the effects of different external aid implementation strategies. These strategies include consistent annual allocation of the same amount over a three-year period, equal distribution over the first two years with no external aid in the third year, and allocating the entire amount only in the first year. The findings reveal that the strategy of allocating external aid equally each year over the three year period results in the lowest total number of disease cases. Additional strategies, including finer stratification into biannual, quarterly, and monthly allocations, highlight the potential for an optimal approach. Specifically, distributing all external aid equally emerges as a strategy associated with the lowest total number of infectious humans. Moreover, distributing all external aid within six, three, or one month results in a higher total number of infectious humans compared to distributing aid within one year. The most significant reduction in the total number of infectious cases occurs with the equal monthly distribution strategy compared to the equal annual, bi-annual, and quarterly distribution strategies.

Simulations of the framework demonstrate the possibility of long transients [57, 72–74]. This phenomenon describes extended periods during which a system takes significant time to reach stability due to complex interactions, feedback loops, or time delays. Understanding long transients is crucial for predicting dynamic system behavior and studying factors influencing transitions. In epidemiology, long transients are relevant for understanding disease dynamics, indicating potential infection reservoirs that require identification and addressing to prevent resurgence.

It should be noted that certain simplifying assumptions have been incorporated into the model framework, potentially influencing outcomes and constraining its applicability. However, the relaxation of these assumptions would introduce increased complexity, rendering the model framework more mathematically intractable. For example, the disease model assumes homogeneity within the population, considering individuals as uniform entities with equal susceptibility and recovery rates. In reality, populations can be heterogeneous, and individual variations in immunity, exposure, and recovery can impact disease dynamics. The model does not account for exposed humans and mosquitoes, which could lead to a delay in the observed dynamics. Also, the model neglects spatial aspects, treating the entire population as a single homogeneous unit. Malaria transmission, however, is influenced by geographical factors, such as mosquito breeding sites and climate, which are crucial for a comprehensive understanding. The framework ignores factors such as mosquito behavior, breeding habitats, and insecticide resistance, which are critical for understanding malaria transmission dynamics. The Solow growth model with a Cobb-Douglas function, insightful as it is in explaining economic growth, encounters limitations. It assumes homogeneous capital and labor, neglecting variations in skills and education. The model also assumes constant returns to scale, a fixed savings rate, and places limited emphasis on technological progress, disregarding the crucial role of technology in sustaining growth. Additionally, it lacks consideration for human capital, assumes full employment, homogeneity of output, and lacks distributional analysis, overlooking income inequality’s real-world implications. While external aid is constant in this study, it should be noted that it can vary as a function of a country’s disease burden. A possible extension of the project includes accounting for skilled and unskilled labor and using the more general constant elasticity of substitution function. Another possible extension includes using an optimal control approach to identify an external aid strategy that will minimize disease prevalence, while maximizing the economic output. Other possible extensions accounting for specific malaria control and mitigation measures and using both malaria data economic data to calibrate the parameters of the model, especially the assumed economic parameters.

In conclusion, the study underscores the intricate interplay between malaria dynamics and economic factors, showcasing bidirectional links between malaria burden and economic development. Hence, the study emphasizes the pivotal role of economics in decision-making for effective disease control. The integrated framework, coupling an epidemiological model of malaria with an economic growth model, provides insights into disease control and highlights the importance of optimizing external aid allocation, particularly favoring strategies with even distribution at short time intervals. The occurrence of bistability, characterized by a backward bifurcation, underscores the challenges of achieving disease elimination, highlights the robustness of elimination states, with the possibility of minor parameter changes triggering significant fluctuations, and the requirement for sustained control measures. The study reveals the potential for long transients, emphasizing the need to address infectious disease reservoirs, as well as the need for extended control measures and continuous monitoring to prevent disease resurgence. Furthermore, the study highlights the nuanced effects of disease and economic parameters on various model outcomes, emphasizing the reciprocal relationship between malaria and per capita yield. In summary, policy recommendations include prioritizing sustained control measures, optimizing aid allocation, and understanding the nuanced inter-dependencies between disease and economic parameters for effective malaria control and prevention.

## Data Availability

All data produced in the present study are available upon reasonable request to the authors

## Acknowledgement

CNN acknowledges the support of the National Science Foundation (Grant Number: DMS #2151870). OP is supported by the National Science Foundation (Grant Number: DMS #2151871), and RZ and HE are supported by the National Science Foundation (Grant Number: DMS #2151872).

## References

[1] World Health Organization (WHO), World malaria report 2021, World Health Organization, Geneva, 2022. URL: https://www.who.int/teams/global-malaria-programme/reports/world-malaria-report-2022.

[2] A. T. Aborode, K. B. David, O. Uwishema, A. L. Nathaniel, J. O. Imisioluwa, S. B. Onigbinde, F. Farooq, Fighting COVID-19 at the expense of malaria in Africa: the consequences and policy options, The American Journal of Tropical Medicine and Hygiene 104 (2021) 26.

[3] G. Bingham, C. Strode, L. Tran, P. T. Khoa, H. P. Jamet, Can piperonyl butoxide enhance the efficacy of pyrethroids against pyrethroid-resistant aedes aegypti?, Tropical Medicine & International Health 16 (2011) 492–500.

[4] S. K. Dadzie, J. Chabi, A. Asafu-Adjaye, O. Owusu-Akrofi, A. Baffoe-Wilmot, K. Malm, C. Bart-Plange, S. Coleman, M. A. Appawu, D. A. Boakye, Evaluation of piperonyl butoxide in enhancing the efficacy of pyrethroid insecticides against resistant anopheles gambiae sl in Ghana, Malaria journal 16 (2017) 1–11.

[5] K. Gleave, N. Lissenden, M. Richardson, L. Choi, H. Ranson, Piperonyl butoxide (PBO) combined with pyrethroids in insecticide-treated nets to prevent malaria in Africa, Cochrane Database of Systematic Reviews (2018).

[6] J. L. Martin, F. W. Mosha, E. Lukole, M. Rowland, J. Todd, J. D. Charlwood, J. F. Mosha, N. Protopopoff, Personal protection with PBO-pyrethroid synergist-treated nets after 2 years of household use against pyrethroid-resistant anopheles in Tanzania, Parasites & Vectors 14 (2021) 1–8.

[7] J. L. Gallup, J. D. Sachs, The economic burden of malaria, The American Journal of Tropical Medicine and Hygiene 64 (2001) 85–96.

[8] J. Sachs, P. Malaney, The economic and social burden of malaria, Nature 415 (2002) 680.

[9] M. T. White, L. Conteh, R. Cibulskis, A. C. Ghani, Costs and cost-effectiveness of malaria control interventions-a systematic review, Malaria journal 10 (2011) 1–14.

[10] S. Tang, D. Feng, R. Wang, B. Ghose, T. Hu, L. Ji, T. Wu, H. Fu, Y. Huang, Z. Feng, Economic burden of malaria inpatients during national malaria elimination programme: estimation of hospitalization cost and its inter-province variation, Malaria journal 16 (2017) 1–10.

[11] F. Mugisha, B. Kouyate, A. Gbangou, R. Sauerborn, Examining out-of-pocket expenditure on health care in Nouna, Burkina Faso: implications for health policy, Tropical Medicine & International Health 7 (2002) 187–196.

[12] J. Nabyonga Orem, F. Mugisha, A. P. Okui, L. Musango, J. M. Kirigia, Health care seeking patterns and determinants of out-of-pocket expenditure for malaria for the children under-five in Uganda, Malaria journal 12 (2013) 1–11.

[13] A. Haakenstad, A. C. Harle, G. Tsakalos, A. E. Micah, T. Tao, M. Anjomshoa, J. Cohen, N. Fullman, S. I. Hay, T. Mestrovic, et al., Tracking spending on malaria by source in 106 countries, 2000–16: an economic modelling study, The Lancet infectious diseases 19 (2019) 703–716.

[14] M. A. Cole, E. Neumayer, The impact of poor health on total factor productivity, The Journal of Development Studies 42 (2006) 918–938.

[15] C. Leighton, R. Foster, Economic impacts of malaria in Kenya and Nigeria, Abt Associates Bethesda, 1993.

[16] D. Fernando, D. De Silva, R. Carter, K. N. Mendis, R. Wickremasinghe, A randomized, double-blind, placebo-controlled, clinical trial of the impact of malaria prevention on the educational attainment of school children, The American journal of tropical medicine and hygiene 74 (2006) 386–393.

[17] R. I. Chima, C. A. Goodman, A. Mills, The economic impact of malaria in Africa: a critical review of the evidence, Health policy 63 (2003) 17–36.

[18] K. Asenso-Okyere, F. A. Asante, J. Tarekegn, K. S. Andam, A review of the economic impact of malaria in agricultural development, Agricultural economics 42 (2011) 293–304.

[19] D. Willis, N. Hamon, Eliminating malaria by 2040 among agricultural households in Africa: potential impact on health, labor productivity, education and gender equality., Gates open research 2 (2018) 33.

[20] D. McCarthy, H. C. Wolf, Y. Wu, The growth costs of malaria, 2000.

[21] F. D. McCarthy, H. C. Wolf, Y. Wu, Malaria and growth, Available at SSRN 629153 (2000).

[22] J. Rosselló, M. Santana-Gallego, W. Awan, Infectious disease risk and international tourism demand, Health policy and planning 32 (2017) 538–548.

[23] P. Malaney, A. Spielman, J. Sachs, The malaria gap, The Intolerable Burden of Malaria II: What’s New, What’s Needed: Supplement to Volume 71 (2) of the American Journal of Tropical Medicine and Hygiene (2004).

[24] S. Fernando, D. Gunawardene, M. Bandara, D. De Silva, R. Carter, K. Mendis, A. Wickremasinghe, The impact of repeated malaria attacks on the school performance of children (2003).

[25] S. D. Fernando, C. Rodrigo, S. Rajapakse, The’hidden’burden of malaria: cognitive impairment following infection, Malaria journal 9 (2010) 1–11.

[26] R. E. Cibulskis, P. Alonso, J. Aponte, M. Aregawi, A. Barrette, L. Bergeron, C. A. Fergus, T. Knox, M. Lynch, E. Patouillard, et al., Malaria: global progress 2000–2015 and future challenges, Infectious diseases of poverty 5 (2016) 1–8.

[27] J.-C. Berthélemy, J. Thuilliez, O. Doumbo, J. Gaudart, Malaria and protective behaviours: is there a malaria trap?, Malaria journal 12 (2013) 1–9.

[28] N. Sarma, E. Patouillard, R. E. Cibulskis, J.-L. Arcand, The economic burden of malaria: revisiting the evidence, The American journal of tropical medicine and hygiene 101 (2019) 1405.

[29] J. Brooke, D. Sridhar, Challenges in tracking global malaria spending, The Lancet Infectious Diseases 19 (2019) 672–673.

[30] World Health Organization, et al., Global health estimates 2016: Disease burden by cause, age, sex, by country and by region, 2000–2016; 2018, Geneva. Recuperado de: https://www.who.int/healthinfo/globalburdendisease/estimates/en/index1.html (2020).

[31] Johns Hopkins Bloomberg School of Public Health, Malaria, Center for Communication Programs, 2022. URL: https://www.malariafreefuture.org/malaria.

[32] R. Ross, The prevention of malaria, John Murray, 1911.

[33] G. Macdonald, The epidemiology and control of malaria., The Epidemiology and Control of Malaria. (1957).

[34] G. A. Ngwa, W. S. Shu, A mathematical model for endemic malaria with variable human and mosquito populations, Mathematical and Computer Modelling 32 (2000) 747–763.

[35] N. Chitnis, J. M. Hyman, J. M. Cushing, Determining important parameters in the spread of malaria through the sensitivity analysis of a mathematical model, Bulletin of Mathematical Biology 70 (2008) 1272–1296.

[36] M. I. Teboh-Ewungkem, G. A. Ngwa, C. N. Ngonghala, Models and proposals for malaria: a review, Mathematical Population Studies 20 (2013) 57–81.

[37] G. A. Ngwa, On the Population Dynamics of the Malaria Vector, Bulletin of Mathematical Biology 68 (8) (2006) 2161–2189.

[38] C. N. Ngonghala, G. A. Ngwa, M. I. Teboh-Ewungkem, Periodic oscillations and backward bifurcation in a model for the dynamics of malaria transmission, Mathematical Biosciences 240 (2012) 45–62.

[39] G. A. Ngwa, T. T. Wankah, M. Y. Fomboh-Nforba, C. N. Ngonghala, M. I. Teboh-Ewungkem, On a reproductive stage-structured model for the population dynamics of the malaria vector, Bulletin of mathematical biology 76 (2014) 2476–2516.

[40] C. N. Ngonghala, M. I. Teboh-Ewungkem, G. A. Ngwa, Persistent oscillations and backward bifurcation in a malaria model with varying human and mosquito populations: implications for control, Journal of Mathematical Biology 70 (2015) 1581–1622.

[41] C. N. Ngonghala, M. I. Teboh-Ewungkem, G. A. Ngwa, Observance of period-doubling bifurcation and chaos in an autonomous ode model for malaria with vector demography, Theoretical Ecology 9 (2016) 337–351.

[42] K. P. Paaijmans, A. F. Read, M. B. Thomas, Understanding the link between malaria risk and climate, Proceedings of the National Academy of Sciences 106 (2009) 13844–13849.

[43] T. M. Lunde, M. N. Bayoh, B. Lindtjørn, How malaria models relate temperature to malaria transmission, Parasites & vectors 6 (2013) 1–10.

[44] E. A. Mordecai, K. P. Paaijmans, L. R. Johnson, C. Balzer, T. Ben-Horin, E. de Moor, A. McNally, S. Pawar, S. J. Ryan, T. C. Smith, et al., Optimal temperature for malaria transmission is dramatically lower than previously predicted, Ecology letters 16 (2013) 22–30.

[45] S. E. Eikenberry, A. B. Gumel, Mathematical modeling of climate change and malaria transmission dynamics: a historical review, Journal of Mathematical Biology 77 (2018) 857–933.

[46] C. N. Ngonghala, The impact of temperature and decay in insecticide-treated net efficacy on malaria prevalence and control, Mathematical Biosciences 355 (2023) 108936.

[47] N. Chitnis, A. Schapira, T. Smith, R. Steketee, Comparing the effectiveness of malaria vector-control interventions through a mathematical model, The American journal of tropical medicine and hygiene 83 (2010) 230–240.

[48] F. B. Agusto, S. Y. Del Valle, K. W. Blayneh, C. N. Ngonghala, M. J. Goncalves, N. Li, R. Zhao, H. Gong, The impact of bed-net use on malaria prevalence, Journal of Theoretical Biology 320 (2013) 58–65.

[49] C. N. Ngonghala, S. Y. Del Valle, R. Zhao, J. Mohammed-Awel, Quantifying the impact of decay in bed-net efficacy on malaria transmission, Journal of Theoretical Biology 363 (2014) 247–261.

[50] C. N. Ngonghala, J. Mohammed-Awel, R. Zhao, O. Prosper, Interplay between insecticide-treated bed-nets and mosquito demography: implications for malaria control, Journal of Theoretical Biology 397 (2016) 179–192.

[51] C. N. Ngonghala, J. Wairimu, J. Adamski, H. Desai, The impact of adaptive mosquito behavior and insecticide-treated nets on malaria prevalence, Journal of Biological Systems 28 (2020) 515–542.

[52] C. N. Ngonghala, Assessing the impact of insecticide-treated nets in the face of insecticide resistance on malaria control, Journal of Theoretical Biology 555 (2022) 111281.

[53] J. Sachs, J. W. McArthur, G. Schmidt-Traub, M. Kruk, C. Bahadur, M. Faye, G. McCord, Ending Africa’s poverty trap, Brookings papers on economic activity 2004 (1) (2004) 117–240.

[54] C. N. Ngonghala, M. M. Pluciński, M. B. Murray, P. E. Farmer, C. B. Barrett, D. C. Keenan, M. H. Bonds, Poverty, disease, and the ecology of complex systems, PLoS Biology 12 (2014) e1001827.

[55] C. N. Ngonghala, G. A. De Leo, M. M. Pascual, D. C. Keenan, A. P. Dobson, M. H. Bonds, General ecological models for human subsistence, health and poverty, Nature Ecology & Evolution 1 (2017) 1153.

[56] M. H. Bonds, A. P. Dobson, D. C. Keenan, Disease ecology, biodiversity, and the latitudinal gradient in income, PLoS Biology 10 (2012) e1001456.

[57] Y. Tao, J. L. Hite, K. D. Lafferty, D. J. Earn, N. Bharti, Transient disease dynamics across ecological scales, Theoretical Ecology (2021) 1–16.

[58] The Global Fund, The Global Fund to Fight AIDS, Tuberculosis and Malaria, Technical Report, Accessed on February 25, 2024. URL: https://www.theglobalfund.org/en/.

[59] President’s Malaia Initiative, U.S. President’s Malaia Initiative, Technical Report, USAID, CDC, Accessed on February 25, 2024. URL: https://www.pmi.gov/.

[60] P. Douglas, C. W. Cobb, A theory of production, The American Economic Review 18 (1928) 139–165.

[61] R. M. Solow, A contribution to the theory of economic growth, The Quarterly Journal of Economics 70 (1956) 65–94.

[62] T. W. Swan, Economic growth and capital accumulation, Economic Record 32 (1956) 334–361.

[63] O. Diekmann, J. A. P. Heesterbeek, J. A. J. Metz, On the definition and computation of the basic reprodcution ratio R0 in models for infectious diseases in heterogeneous populations, Journal of Mathematical Biology 28 (1990) 365–382.

[64] P. Van den Driessche, J. Watmough, Reproduction numbers and sub-threshold endemic equilibria for compartmental models of disease transmission, Mathematical Biosciences 180 (2002) 29–48.

[65] P. M. Romer, Endogenous technological change, Journal of political Economy 98 (1990) S71–S102.

[66] N. G. Mankiw, D. Romer, D. N. Weil, A contribution to the empirics of economic growth, The quarterly journal of economics 107 (1992) 407–437.

[67] A. Gumel, Causes of backward bifurcations in some epidemiological models, Journal of Mathematical Analysis and Applications 395 (2012) 355–365.

[68] C. Castillo-Chavez, B. Song, Dynamical models of tuberculosis and their applications, Math. Biosci. Eng 1 (2004) 361–404.

[69] M. M. Pluciński, C. N. Ngonghala, M. H. Bonds, Health safety nets can break cycles of poverty and disease: a stochastic ecological model, Journal of The Royal Society Interface 8 (2011) 1796–1803.

[70] A. Garchitorena, S. H. Sokolow, B. Roche, C. N. Ngonghala, M. Jocque, A. Lund, M. Barry, E. A. Mordecai, G. C. Daily, J. H. Jones, J. R. Andrews, E. Bendavid, S. P. Luby, A. D. LaBeaud, K. Seetah, J. F. Guégan, M. H. Bonds, G. A. De Leo, Disease ecology, health and the environment: a framework to account for ecological and socio-economic drivers in the control of neglected tropical diseases, Philosophical Transactions of the Royal Society B: Biological Sciences 372 (2017) 20160128.

[71] T. Jezek, Country size bias in global health: cross-country comparison of malaria policy and foreign aid, Global Health Research and Policy 6 (2021) 1–12.

[72] A. Hastings, Transients: the key to long-term ecological understanding?, Trends in ecology & evolution 19 (2004) 39–45.

[73] A. Morozov, K. Abbott, K. Cuddington, T. Francis, G. Gellner, A. Hastings, Y.-C. Lai, S. Petrovskii, K. Scranton, M. L. Zeeman, Long transients in ecology: Theory and applications, Physics of Life Reviews 32 (2020) 1–40.

[74] T. B. Francis, K. C. Abbott, K. Cuddington, G. Gellner, A. Hastings, Y.-C. Lai, A. Morozov, S. Petrovskii, M. L. Zeeman, Management implications of long transients in ecological systems, Nature Ecology & Evolution 5 (2021) 285–294.

