## Supplementary Material for "Modeling the synergistic interplay between malaria dynamics and economic growth" (from another version of this preprint: version 1, 2023-12-10)

### Supplementary Information (SI)

#### S1. Next generation operator approach

The Next Generation operator method developed by [1, 2] is a powerful mathematical tool commonly used in epidemiology to analyze and predict the transmission dynamics of infectious diseases. It involves the use of matrices to model disease spread and assess key parameters like the basic reproduction number ($R_0$), which helps determine the disease's potential for sustained transmission in a population. This method is particularly valuable for understanding disease control measures and informing public health interventions. We show that the conditions for the next generationj operator approach are satisfied for the model (2.1) in the main text.

**Theorem S1.1.** *Assume $\alpha \in (0,1)$. If $y_E > 0$, all conditions of the next generation approach (A1–A5 below) are satisfied if the state variable, $k$, representing capital is treated as a disease-free state. If $y_E = 0$, all conditions of the next generation matrix approach are satisfied when the linearization is about the locally asymptotically stable disease-free equilibrium; that is, the DFE with $k^0 = k_2^0 > 0$. In either case, $R_0^2 = \frac{p_{vh}\beta_v(y^0)}{\mu_v(y^0)} \cdot \frac{p_{hv}\beta_v(y^0)}{\delta_h+\gamma(y^0)+\mu_h(y^0)} \left(1 + \frac{\theta\gamma_h(y^0)}{\rho_h+\mu_h(y^0)}\right) \frac{S_v^0}{S_h^0}$, where $S_h^0 = \frac{\Lambda_{h0}}{\mu_h}$, $S_v^0 = \frac{\Lambda_{v0}(y^0)}{\mu_v(y^0)}$, $k^0$ is the unique positive solution of the equilibrium equation $-k\,(\sigma) - r_0\,(c-1)\left(A^{1-\alpha}k^{\alpha} + y_E\right) = 0$, and $y^0 = A^{1-\alpha}k^{\alpha} + y_E$.*

*Proof.* Consider the ordering of the state variables $\vec{x} = (x_1, x_2, x_3, x_4, x_5, x_6, x_7) = (S_h, S_v, N_h, k, I_h, R_h, I_v)$. The disease free states are $X_s = (x_1, x_2, x_3, x_4) = (S_h, S_v, N_h, k)$ and the diseased states are $(x_5, x_6, x_7) = (I_h, R_h, I_v)$. We will show that there is a construction of the vectors $\mathcal{F}$ and $\mathcal{V}$ such that $\vec{x}' = \mathcal{F} - \mathcal{V}$ satisfies the following assumptions (A1–A5) of the Next Generation Matrix approach.

- (A1) if $x \geq 0$, then $\mathcal{F}_i, \mathcal{V}_i^+$, and $\mathcal{V}_i^-$ are nonnegative for $i = 1, \ldots, 7$.
- (A2) if $x_i = 0$ then $V_i^- = 0$. In particular, if $x \in X_s$, then $V_i^- = 0$ for $i = 5, 6, 7$.
- (A3) $F_i = 0$ for $i < 5$.
- (A4) if $x \in X_s$ then $\mathcal{F}_i(x) = 0$ and $\mathcal{V}_i^+(x) = 0$ for $i = 5, 6, 7$.
- (A5) if $\mathcal{F}(x)$ is set to zero, then all eigenvalues of the Jacobian $Df(x_0)$ have negative real part (that is, the disease-free equilibrium is locally asymptotically stable in the absence of new infection).

Define $\mathcal{F}$ to be a vector of the new infections terms, and $\mathcal{V}$ to be a vector of the remaining transitions. Then,

$$
\mathcal{F} = \begin{bmatrix} 0 \\ 0 \\ 0 \\ 0 \\ \frac{I_v S_h p_{vh} \beta_v y}{N_h} \\ 0 \\ \frac{S_v p_{hv} (I_h + R_h \theta) \beta_v y}{N_h} \end{bmatrix} \tag{S1}
$$

and

$$
\mathcal{V} = \begin{bmatrix} -\Lambda_h - R_h \rho_h + S_h \left( \frac{I_v p_{vh} \beta_v y}{N_h} + \mu_h \right) \\ S_v \left( \mu_v y + \frac{p_{hv} (I_h + R_h \theta) \beta_v y}{N_h} \right) - \Lambda_v y \\ I_h \delta_h - \Lambda_h + N_h \mu_h \\ k \left( \sigma + \frac{-I_h \delta_h + \Lambda_h - N_h \mu_h}{N_h} \right) - r_0 y \left( -\frac{I_h \xi}{N_h} - c + 1 \right) \\ I_h \left( \delta_h + \mu_h + \gamma y \right) \\ -I_h \gamma y + R_h \left( \mu_h + \rho_h \right) \\ I_v \mu_v y \end{bmatrix}, \tag{S2}
$$

recalling that $y = \left( k^{\alpha} \left( \frac{A(I_h(1-\epsilon) + R_h + S_h)}{N_h} \right)^{1-\alpha} + y_E \right)$.

We can write $\mathcal{V} = \mathcal{V}^+ - \mathcal{V}^-$, where

$$
V^+ = \begin{bmatrix} S_h \left( \frac{I_v p_{vh} \beta_v y}{N_h} + \mu_h \right) \\ S_v \left( \mu_v y + \frac{p_{hv} (I_h + R_h \theta) \beta_v y}{N_h} \right) \\ I_h \delta_h + N_h \mu_h \\ k \left( \sigma + \frac{\Lambda_h}{N_h} \right) + r_0 y \left( \frac{I_h \xi}{N_h} + c \right) \\ I_h \left( \delta_h + \mu_h + \gamma y \right) \\ R_h \left( \mu_h + \rho_h \right) \\ I_v \mu_v y \end{bmatrix} \text{ and } V^- = \begin{bmatrix} \Lambda_h + R_h \rho_h \\ \Lambda_v y \\ \Lambda_h \\ k \left( \frac{I_h \delta_h + N_h \mu_h}{N_h} \right) + r_0 y \\ 0 \\ I_h \gamma y \\ 0 \end{bmatrix}
$$

The conditions (A1)-(A4) are immediately satisfied by this construction of $\mathcal{F}$ and $\mathcal{V}$. This leaves us with only condition (A5) to verify. This condition will be satisfied if the Jacobian of $-\mathcal{V}$ evaluated at the disease-free equilibrium has all eigenvalues in the left half of the complex plane. It should be noted that the Jacobians of $\mathcal{F}$ and $\mathcal{V}$ are

$$
F = \begin{bmatrix} 0 & 0 & 0 & 0 & 0 & 0 & f_{17} \\ 0 & 0 & 0 & 0 & f_{25} & f_{26} & 0 \\ 0 & 0 & 0 & 0 & 0 & 0 & 0 \\ 0 & 0 & 0 & 0 & 0 & 0 & 0 \\ 0 & 0 & 0 & 0 & 0 & 0 & f_{57} \\ 0 & 0 & 0 & 0 & 0 & 0 & 0 \\ 0 & 0 & 0 & 0 & f_{75} & f_{76} & 0 \end{bmatrix} \text{ and } V = \begin{bmatrix} v_{11} & 0 & 0 & 0 & 0 & v_{16} & 0 \\ v_{21} & v_{22} & v_{23} & v_{24} & v_{25} & v_{26} & 0 \\ 0 & 0 & v_{33} & 0 & v_{35} & 0 & 0 \\ v_{41} & 0 & v_{43} & v_{44} & v_{45} & v_{46} & 0 \\ 0 & 0 & 0 & 0 & v_{55} & 0 & 0 \\ 0 & 0 & 0 & 0 & v_{65} & v_{66} & 0 \\ 0 & 0 & 0 & 0 & 0 & 0 & v_{77} \end{bmatrix},
$$

respectively, where

$$
f_{17} =, \quad f_{25} =, \quad f_{26} =,
$$

$$f_{57} = p_{vh}\beta_v\big(A^{1-\alpha}(k^0)^\alpha + y_E\big), f_{75} = \frac{S_v^0 p_{hv}\beta_v\big(A^{1-\alpha}(k^0)^\alpha + y_E\big)}{S_h^0}, f_{76} = \frac{S_v^0 p_{hv}\theta\beta_v\big(A^{1-\alpha}(k^0)^\alpha + y_E\big)}{S_h^0}.$$

The eigenvalues of $V$ are $v_{ii}$ for $i = 1, \ldots, 7$. If each of these eigenvalues has a positive real part, then the disease-free equilibrium is locally asymptotically stable, and (A5) is satisfied. The eigenvalues are:

$$v_{11} = \mu_h, \quad v_{22} = \mu_v\big(A^{1-\alpha}(k^0)^\alpha + y_E\big), \quad v_{33} = \mu_h, \quad v_{44} = \sigma - \frac{A^{1-\alpha}\alpha(k^0)^\alpha r_0\,(1-c)}{k^0}$$
$$v_{55} = \delta_h + \mu_h + \gamma\big(A^{1-\alpha}(k^0)^\alpha + y_E\big), \quad v_{66} = \mu_h + \rho_h, \quad v_{77} = \mu_v\big(A^{1-\alpha}(k^0)^\alpha + y_E\big).$$

The only eigenvalue with a potentially ambiguous sign is $v_{44}$. However, recall that $k^0$ is the unique positive solution of the equilibrium equation $-k\sigma - r_0\,(c-1)\,\big(A^{1-\alpha}k^\alpha + y_E\big) = 0$. Solving for $\sigma$ and evaluating at $k = k^0$ yields $\sigma = \frac{r_0(1-c)\big(A^{1-\alpha}(k^0)^\alpha + y_E\big)}{k^0}$. Substituting this expression into $v_{44}$ yields $v_{44} = \frac{r_0(1-c)}{k^0}\big[(1-\alpha)A^{1-\alpha}(k^0)^\alpha + y_E\big]$, which is positive given the parameter constraints $\alpha \in [0,1]$ and $c \in [0,1)$. Therefore, (A5) is satisfied.

We now define $\rho(NGM)$ to be the spectral radius of the so called next generation matrix $NGM = FV^{-1}$, where

$$NGM = \begin{bmatrix} 0 & 0 & 0 & 0 & 0 & 0 & \frac{f_{17}}{v_{77}} \\ 0 & 0 & 0 & 0 & \frac{f_{25}}{v_{55}} - \frac{f_{26}v_{65}}{v_{55}v_{66}} & \frac{f_{26}}{v_{66}} & 0 \\ 0 & 0 & 0 & 0 & 0 & 0 & 0 \\ 0 & 0 & 0 & 0 & 0 & 0 & 0 \\ 0 & 0 & 0 & 0 & 0 & 0 & \frac{f_{57}}{v_{77}} \\ 0 & 0 & 0 & 0 & 0 & 0 & 0 \\ 0 & 0 & 0 & 0 & \frac{f_{75}}{v_{55}} - \frac{f_{76}v_{65}}{v_{55}v_{66}} & \frac{f_{76}}{v_{66}} & 0 \end{bmatrix}, \tag{S3}$$

which has spectral radius $\rho(NGM) - \sqrt{\frac{f_{57}(f_{75}v_{66} - f_{76}v_{65})}{v_{55}v_{66}v_{77}}}$, and

$$\begin{aligned} f_{57} &= p_{vh}\beta_v\big(A^{1-\alpha}(k^0)^\alpha + y_E\big) \\ f_{75} &= \frac{S_v^0 p_{hv}\beta_v\big(A^{1-\alpha}(k^0)^\alpha + y_E\big)}{S_h^0} \\ f_{76} &= \frac{S_v^0 p_{hv}\theta\beta_v\big(A^{1-\alpha}(k^0)^\alpha + y_E\big)}{S_h^0} \\ v_{65} &= -\gamma\big(A^{1-\alpha}(k^0)^\alpha + y_E\big). \end{aligned}$$

According to the next generation matrix method, if assumptions (A1)-(A5) hold, the disease-free equilibrium is locally asymptotically stable if $\rho(NGM) < 1$, and unstable if $\rho(NGM) > 1$. We define $R_0 = (\rho(NGM))^2$, which completes the proof.

□

#### S2. Baseline parameter values

Table S1: Parameters of model (2.1). The epidemiological baseline parameter values are drawn from [3–5].

| Parameter | Description | Dimension | Baseline value |
|---|---|---|---|
| $\Lambda_h$ | Human recruitment rate | Humans $\times$ day$^{-1}$ | $1500/(55 \times 365)$ |
| $\Lambda_{v0}$ | Mosquito recruitment rate | Mosquitoes $\times$ day$^{-1}$ | $10^4/14$ |
| $\mu_h$ | Human natural death rate | day$^{-1}$ | $1/(55 \times 365)$ |
| $\mu_{v0}$ | Mosquito death rate | day$^{-1}$ | $1/14$ |
| $\mu_{v1}$ | Additional mosquito death rate | day$^{-1}$ | $1/14$ |
| $\delta_h$ | Human disease-induced death rate | day$^{-1}$ | $32.9/(365\times1000)$ |
| $\gamma_0$ | Rate at which infectious humans recover | day$^{-1}$ | $1/180$ |
| $\gamma_1$ | Additional rate at which infectious humans recover | day$^{-1}$ | $1/14 - 1/180$ |
| $\rho_h$ | Rate at which immune humans lose immunity | day$^{-1}$ | $1/(90)$ |
| $\beta_{v0}$ | Mosquito biting rate | day$^{-1}$ | $9.5/10$ |
| $p_{vh}$ | Disease transmission probability from infectious mosquitoes to humans | Dimensionless | $22/1000$ |
| $p_{hv}$ | Disease transmission probability from infectious humans to mosquitoes | Dimensionless | $48/100$ |
| $\theta p_{hv}$ | Disease transmission probability from immune humans to mosquitoes | Dimensionless | $48/1000$ |
| $A$ | Labor efficiency | | 1 |
| $\varepsilon$ | Reduce productivity factor due to illness | Dimensionless | 0.5 |
| $c$ | Consumed portion of the yield | Dimensionless | 0.4 |
| $\xi$ | Medical cost reduction factor due to illness | Dimensionless | 0.5 |
| $\sigma$ | Rate that yield depreciates | day$^{-1}$ | 0.001 |
| $r_0$ | Rate that unconsumed yield accumulates | day$^{-1}$ | 2*$\sigma$ |
| $y_E$ | External yield | | 1 |
| $\alpha$ | Captical share | | .5 |
| $\mu_{vy}$ | Saturation factor | | 10 |
| $\gamma_y$ | Saturation factor | | 10 |
| $\beta_y$ | Saturation factor | | 10 |
| $\Lambda_{vy}$ | Saturation factor | | 10 |

Table S2: Possible number of positive equilibrium points of the model (2.1).

| $B_2$ | $B_1$ | $B_0$ | Number of equilibira |
|---|---|---|---|
| Always positive | Positive when $M \leq 1$ | Positive when $R_0 \leq 1$ | 0 |
| Always positive | Positive when $M \leq 1$ | Negative when $R_0 > 1$ | 1 |
| Always positive | Negative when $M > 1$ | Negative when $R_0 > 1$ | 1 |
| Always positive | Negative when $M > 1$ | Positive when $R_0 < 1$ | 2 |

#### S3. More simulations

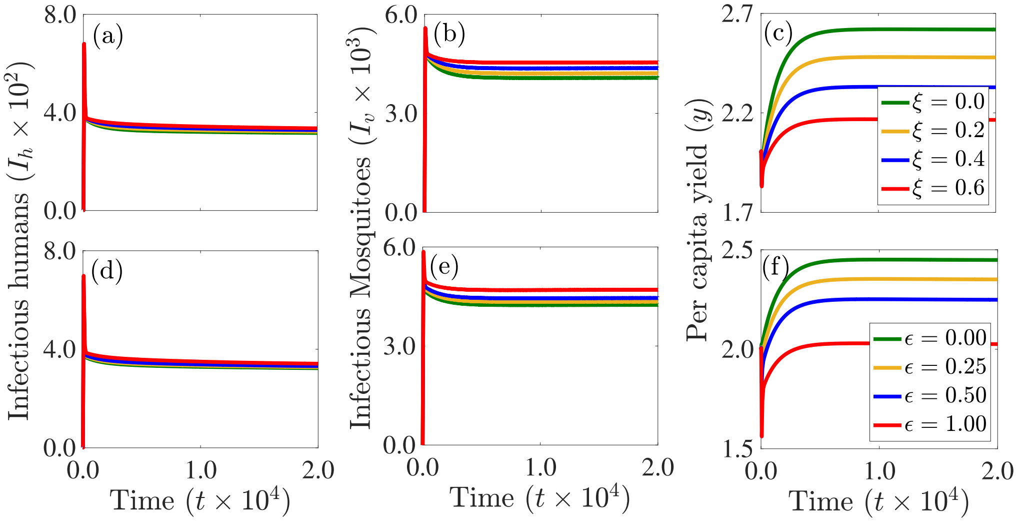


Figure S1: Simulations of the model (2.1) depicting the long term dynamics of the infectious human population ($I_h$: (a) and (d)), infectious mosquito population ($I_v$: (b) and (e)), and per capita yield ($y$: (c) and (f)) for different values of the malaria-related medical costs parameter ($\xi$: (a)-(c)) and the adjustment factor for decreased productivity associated with clinical malaria ($\varepsilon$: (d)-(f)). The initial condition used for the simulations is $(S_h(0), I_h(0), R_h(0), S_v(0), I_v(0), k(0)) = (999, 1, 0, 10000, 0, 1)$, while the values of the other parameters are presented in Table S1.

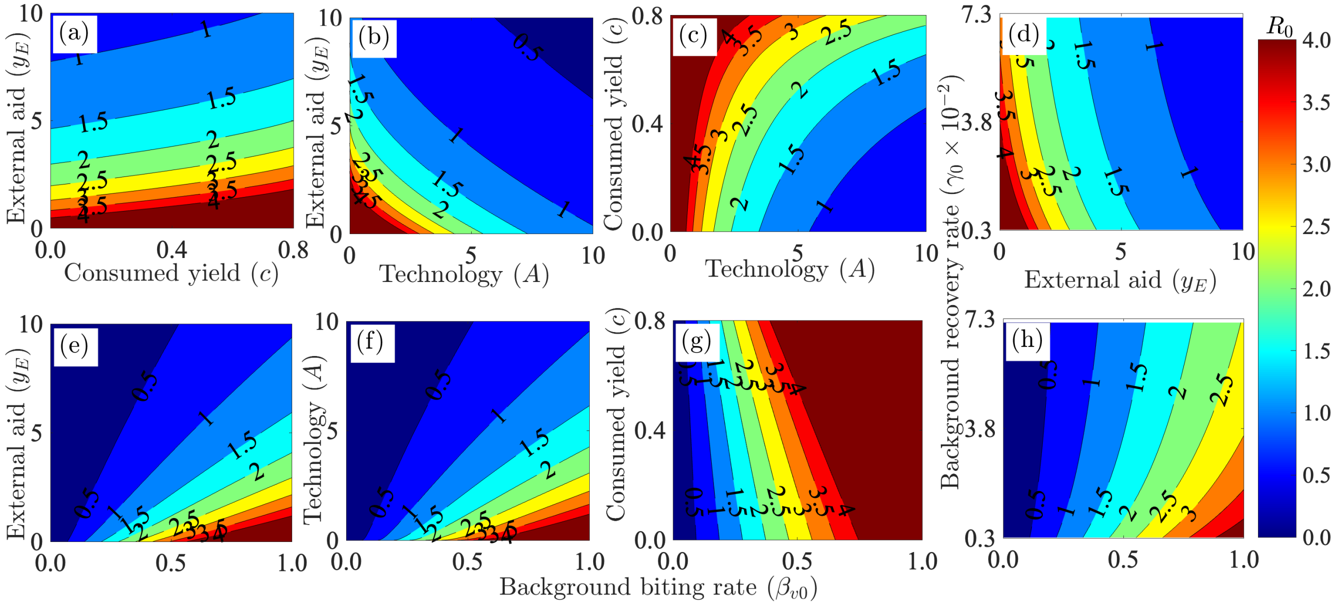


Figure S2: heat maps of the basic reproduction number ($R_0$) of the model (2.1) as a function of (a) the fraction of the yield consumed ($c$) and external aid ($y_E$), (b) technological progress or labor efficiency ($A$) and external aid ($y_E$), (c) technological progress and the fraction of the yield consumed, (d) external aid and background recovery rate ($\gamma_0$), (e) background mosquito biting rate ($\beta_{v0}$) and external aid, (f) background mosquito biting rate and technological progress, (g) background mosquito biting rate and the fraction of the yield consumed, and (h) background mosquito biting rate and the background recovery rate. With the exception of these varied parameters, the other parameters are held at their baseline values given in Table S1.

#### References


[1] O. Diekmann, J. A. P. Heesterbeek, J. A. J. Metz, On the definition and computation of the basic reprodcution ratio $R_0$ in models for infectious diseases in heterogeneous populations, Journal of Mathematical Biology 28 (1990) 365–382.

[2] P. Van den Driessche, J. Watmough, Reproduction numbers and sub-threshold endemic equilibria for compartmental models of disease transmission, Mathematical Biosciences 180 (2002) 29–48.

[3] N. Chitnis, J. M. Cushing, J. M. Hyman, Bifurcation analysis of a mathematical model for malaria transmission, SIAM Journal of Applied Mathematics 67 (1) (2006) 24–45.

[4] C. N. Ngonghala, S. Y. Del Valle, R. Zhao, J. Mohammed-Awel, Quantifying the impact of decay in bed-net efficacy on malaria transmission, Journal of Theoretical Biology 363 (2014) 247–261.

[5] F. B. Agusto, S. Y. D. Valle, K. W. Blayneh, C. N. Ngonghala, M. J. Goncalves, N. Li, R. Zhao, H. Gong, The impact of bed-net use on malaria prevalence, Journal of Theoretical Biology 320 (2013) 58–65.
